# Digital solutions supporting the quality of life of European cancer patients and their caregivers: a systematic literature review

**DOI:** 10.1101/2024.06.18.24309065

**Authors:** Camilla Ancona, Emanuele Caroppo, Pietro De Lellis

## Abstract

**Purpose:** we investigate (a) the diffusion of digital solutions supporting the quality of life in cancer patients and their caregivers across cancer types and EU countries, (b) the key thematic areas on which they focus, and (c) their effectiveness in improving the quality of life with respect to traditional healthcare.

**Methods:** We searched articles from Embase, Scopus and PubMed in the last decade, and assessed their quality according to mixed methods appraisal tool. We compared the effectiveness of such tools and discussed the main gaps that emerged.

**Results:** 49 studies were included (31 quantitative randomized control trials, 9 quantitative non-randomized, 4 quantitative descriptive, 3 qualitative, and 2 mixed-methods). We observed a prevalence of studies from the Netherlands and Germany, and breast cancer patients are the most targeted by single-cancer type interventions. The key areas of interventions for e-health solutions are psychophysical well-being, management of physical distress, remote monitoring of vitals and symptoms, and empowerment and self-efficacy. The effectiveness of digital solutions is typically higher than traditional healthcare, especially for solutions focusing on psychosocial well-being.

**Conclusions:** This review showed a growing interest in digital solutions aimed at making the life of cancer patients and their caregivers easier, and their healthcare more patient-centered. The effectiveness of such interventions varies, but all the solutions are well accepted among the participants. Our findings provide evidence of the untapped potential of these digital tools, and of the need for their integration in the daily routine of cancer patients and their caregivers.

## 1. Introduction

Cancer is a pervasive health challenge worldwide, affecting millions of individuals of all ages and sociodemographic backgrounds. Across the 27 EU Member States (EU27), cancer incidence rates are significant, with approximately 2.74 million new cases diagnosed in 2022^1^, with the prevalent cancer sites comprising the breast, prostate, colorectum, and lung, collectively constituting 50% of all incident cancer cases. Thanks to the medicine advances, in the last decade there has been a notable 10% decline in cancer mortality within the EU27, even though there are noticeable disparities in the estimated five-year survival probabilities among EU countries, with Central and Eastern European countries showing lower rates, whereas Western European and Nordic countries consistently manifesting top quintile survival rates.

Intranational differences in cancer mortality rates are also present, reaching up to a 37% variability across distinct regions, underlining the potential for targeted interventions to integrate existing healthcare tools country-wise and mitigate regional disparities. A comprehensive national cancer registry encompassing the entire population is active in 23 out of the 27 EU member states. Among these, only four countries (Spain, Italy, Romania, and France) maintain regional registries spanning varying proportions of their respective populations, while Hungary, Luxemburg, Cyprus and Greece are lacking a population-based cancer registry infrastructure.

Initiatives aimed towards homogenizing standards and favouring interoperability across databases would facilitate the merging of cancer registries and national screening datasets, thereby fostering improved surveillance of cancer prevalence and enhancing cancer care provision. Of notable importance is the facilitation of sociodemographic data linkage with cancer registries, enabling the monitoring of cancer-related inequities and the formulation of targeted policy interventions. National cancer mitigation plans are operational in 23 out of the 27 EU nations, with a strong focus on prevention, screening, and quality of cancer care, but not specifically on cancer network infrastructure, digitalization, and health information systems, which are comparatively less prioritized.

While survival rates have improved over time, the burden of cancer treatment on patients and their caregivers is substantial, encompassing a spectrum of physical symptoms, emotional distress, and practical challenges. Common symptoms experienced by cancer patients include pain, fatigue, nausea, and psychological distress, which can profoundly affect their quality of life (QoL) and functional capacity^2^. Furthermore, the aftermath of cancer treatment may bring about long-term health issues, including chronic conditions, cognitive impairments, and psychosocial difficulties, which necessitate ongoing support and management. Whether diagnosed in childhood, adolescence, or adulthood, the impact of cancer reverberates throughout every aspect of their lives, influencing not only their physical well-being but also their emotional resilience and social connections. Also, informal cancer caregivers face a significant burden, encompassing emotional, physical and financial challenges.

Anxiety, solitude, fear of the future are shared emotions up to the prognosis; fatigue, stress and being overwhelmed by caregiving tasks such as managing symptoms, administering medication, provide daily support; financially, many have to significantly reduce their working hours and undertake high costs of medical care.

Assisting the loved ones who have been diagnosed with cancer constitutes a tricky trade-off for caregivers; a balance has to be reached between providing all the required support and not to neglect their own needs (Services, s.d.).

Traditional face-to-face interventions have been key in addressing the complex needs of cancer patients and survivors. However, accessing these services can be hindered by various barriers, such as geographic distance, long waiting lists, time constraints, and stigma surrounding oncological health care^3,4^. In response to these challenges, digital health interventions have emerged as a promising novelty to provide accessible support to individuals affected by cancer^5–7^.

Digital health interventions encompass a broad spectrum of technologies, as illustrated in Figure 1. The existing spectrum of digital tools in healthcare., including mobile health (mHealth) and electronic health (eHealth) tools, designed to deliver health-related services and interventions remotely by means of mobile devices and web-based platforms^8–10^. These interventions offer the potential to improve medication adherence, self-management, and psychosocial well-being across the cancer continuum, from diagnosis to survivorship. Relieving inconvenient side effects, remote monitoring vitals, simplifying the check-up procedures, reducing anxiety, depression, and solitude, promoting the screening treatments, are only few of the main goals that digital tools are aimed to.

**Figure 1.**
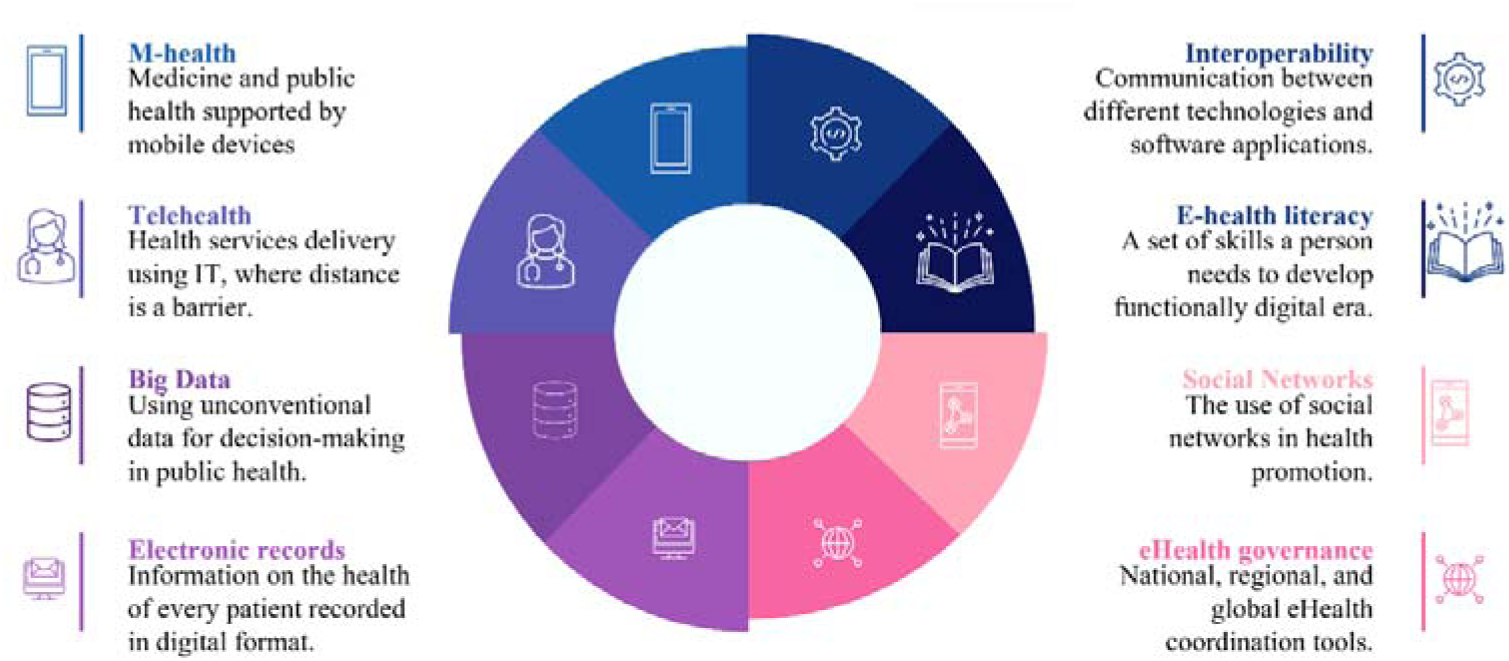
The existing spectrum of digital tools in healthcare.

With the widespread availability of mobile devices and internet connectivity, digital health interventions have become increasingly accessible to individuals of all ages and educational backgrounds. Whether accessed via smartphones, tablets, or computers, these interventions offer a convenient and flexible means of delivering support and resources that can be tailored to the diverse needs of cancer patients and survivors. Previous reviews have often focused on specific cancer types, age groups, or stages of oncological illness^5,8–15^. Despite the growing body of evidence supporting the feasibility, acceptability, and efficacy of digital health interventions in oncology, there remains a need for a comprehensive synthesis of the existing literature, particularly across EU countries.

This systematic literature review aims to address this gap by comprehensively examining the landscape of digital interventions targeting various dimensions of cancer care, including symptom management and monitoring, psychosocial support, all aimed to improve the quality of life of oncological patients and their caregivers, across different cancer types and EU countries. By synthesizing the current evidence and critically appraising intervention usability, effectiveness and rate of adherence, this review seeks to inform the development and implementation of digital health interventions tailored to the diverse needs of individuals affected by cancer.

Understanding the acceptability and feasibility of these interventions is crucial for their successful design and integration into routine cancer care practice.

Following the PICO framework to define our research questions, this systematic review aims to provide insights into the potential of digital health interventions to support cancer patients and their caregivers across the EU countries, and to compare their effectiveness with respect to traditional healthcare in terms of quality of life, see the schematic in Figure 2.

**Figure 2.**
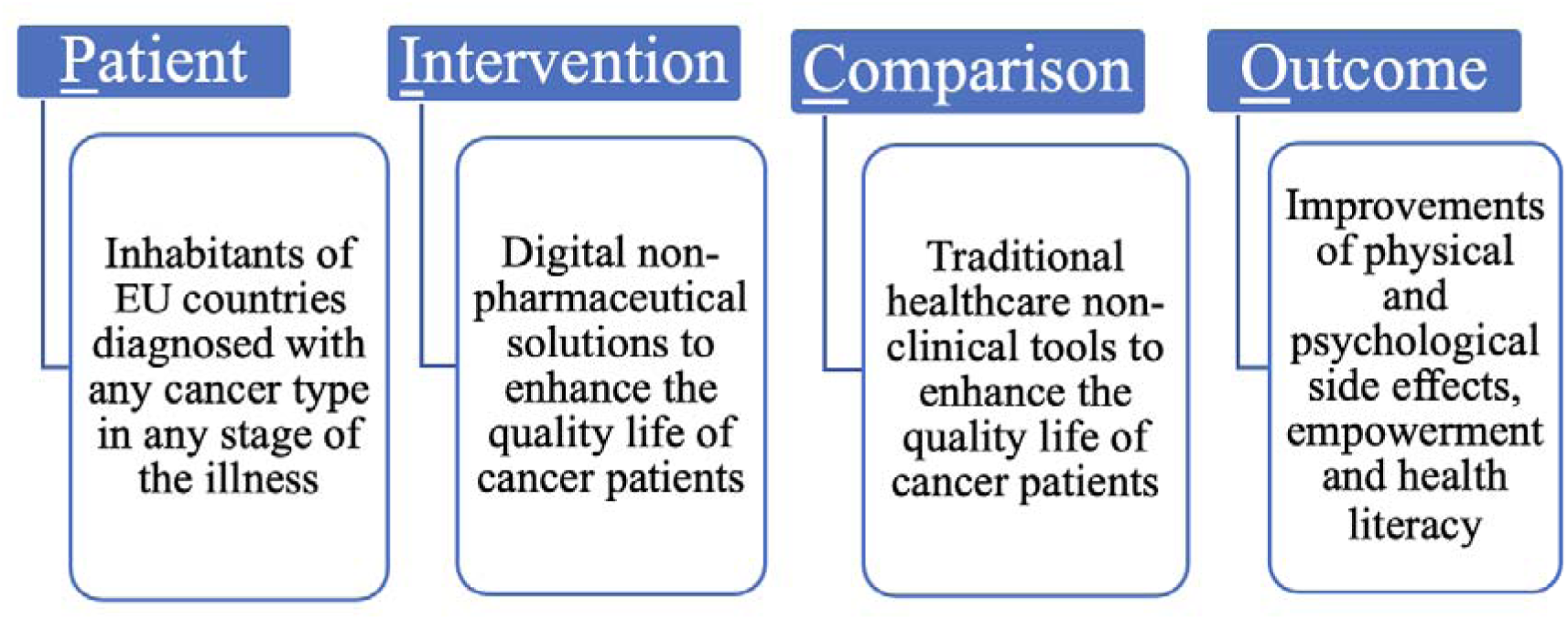
PICO framework for study design.

The research questions that we strive to address are

RQ1) Have digital solutions been uniformly suggested and examined across all cancer types and EU countries?
RQ2) What key areas do digital solutions focus on to enhance the quality of life for cancer patients and their caregivers?
RQ3) What is the effectiveness of the digital solutions in the EU compared to traditional healthcare practices?

The ultimate goal of this review is to aid the progress of supportive care strategies for cancer patients and their caregivers by identifying existing interventions in the literature and highlighting the potential of such initiatives, thereby fostering the integration and cooperation among the EU countries.

## 2. Methods

This systematic review was conducted in accordance with the Preferred Reporting Items for Systematic Reviews and Meta-Analysis (PRISMA) statement guidelines. Meta-analysis was considered unfeasible due to the heterogeneity in study types, methodologies and outcome’s variables reported. Results have been then summarized in tabular form reporting authors and date of publication, country where the study was undertaken, the aim of the digital intervention, the study design, the sample size of the patients included in the study, the type of intervention, the observed feasibility/ usability/ adherence, the primary outcomes and results of the study, see Table 6. The systematic review protocol is registered with the International Prospective Register of Systematic Reviews (PROSPERO) database (CRD42024529592).

### 2.1. Search strategy

We performed a comprehensive search of the literature to identify peer-reviewed journal articles that included the use of electronic interventions in the European Union for people undergoing cancer treatment, cancer survivors or their caregivers. The search was conducted at the end of 2023 and repeated in January 2024 via three electronic databases: PubMed, Embase and Scopus. Other studies have been added among the papers cited in feasibility studies or protocols that have been excluded by means of a snowballing technique. We designed the search combining words related to three main themes: subjects, scopes, and methods. Search terms were merged thanks to Boolean operators, with the final query including the following key words: cancer, oncological, caregiver, support, care, quality of life, improve*, effect*, well*, literacy, efficacy, telehealth, e-health, nonclinical, digital, e-mental, e-solutions, e-support, web-based, e-interventions, app, website, online, AI, wearable, remote, smart, mobile, virtual, technology, applications. Medical Subject Headings (MeSH), or equivalent terms, were used, as well as plural variations of the keywords.

### 2.2 Eligibility Criteria

The criteria guiding the inclusion of articles in this systematic literature review are as follows: I1) Peer-reviewed articles: eligibility was given to studies subjected to peer review processes to ensure the integrity and scientific soundness of the included research endeavors.

I2) Primary studies: reviews, commentaries, protocols, opinion papers, and editorials were excluded, with a focus maintained solely on primary research contributions. For the protocols, we opted to include the primary study that followed, if any.

I3) Language: articles written in the English language were considered for screening, guaranteeing uniformity for analysis, and facilitating a coherent interpretation and synthesis of the findings.

I4) Publication timeframe: articles published in the last decade, that is, from 2014 to 2024, thereby enabling a comprehensive exploration of contemporary developments in digital non-pharmaceutical interventions of cancer care.

I5) Population: we focus on the inhabitants of EU countries diagnosed with cancer across all stages of treatment alongside cancer survivors, and their caregivers. This criterion enables us to provide a wide perspective on the current initiatives across the EU countries.

Conversely, the criteria for the exclusion from the review were defined as follows:

E1) Clinical technology-based methods: studies predominantly oriented towards clinical applications, including tumor spreading management, treatment decision support systems for practitioners, and enhancements in diagnostic imaging were excluded. These areas, being clinical in nature, fall outside the scope of this work and should be handled by specialized medical professionals. This choice underlines the review’s focus on patient-centric interventions rather than clinical or therapeutic methods.

E2) Geographical scope: articles that included inhabitants of countries outside the European Union (EU27) countries were excluded, ensuring alignment with the review’s geographic focus, and enhancing the relevance and applicability of the synthesized findings within the EU healthcare framework, in line with the objectives of the Erasmus+ project “EHealth4Cancer”, from which the concept of this work originated^16^.

E3) Accessibility: articles lacking full-text availability were excluded to safeguard against incomplete or imprecise reports of their findings.

As for the eligible outcomes, we included studies proposing digital solutions, ranging from wearable devices to web-based platforms and apps, aiming at enhancing the health-related quality of life of the patients and their caregivers, encompassing either the physical and psychological well-being, patients’ self-efficacy in managing their healthcare, and the ability to monitor their symptoms and provide valuable information for practitioners.

### 2.3 Data selection

All the authors (C.A., P.D.L and E.C.) designed the search strategy and query and conducted the literature search. Next, C.A. and P.D.L. independently screened the articles sequentially by title, abstract and then full text to determine eligibility based on the specified inclusion/exclusion criteria; disagreements were resolved by a third reviewer (E.C.). Then, they included their decision in a collection of the reference software manager Zotero. They were blinded to each other’s’ decisions during the selection phase. Disagreements have been resolved by consulting a third researcher (E.C.). Regarding the data extraction, all researchers agreed on collecting information about authors’ name, publication year, country, study design, diagnosis, the number of participants of each publication, the country where the study has been conducted, the type of digital intervention, characteristics of participants (i.e., age, gender, and stage of treatment), feasibility, participation and adherence measures and rates, primary outcome measures, and intervention duration. All these data have been stored in excel spreadsheets and some notes and comments have been added in the shared Zotero folder. As for the extraction phase, two authors (C.A. and P.D.L) independently extracted data and disagreements have been resolved by consulting the third author (E.C.). A final review of the full-text articles was conducted by a third researcher (E.C.). The original inclusion and exclusion criteria as published in the PROSPERO registered protocol were followed accurately. Reasons for exclusion of full-text papers were documented in the PRISMA study flowchart depicted in Figure.

### 2.4 Critical appraisal

The analysis was independently assessed by C.A. and P.D.L, then E.C. checked the results for consistency. We did not limit the search to a specific type of study as we wanted to investigate the state-of-the-art regarding the digital solutions for cancer patients and their caregivers in a comprehensive way. Thus, we included both qualitative and quantitative studies to gather both qualitative perceptions and quantitative outcomes on the effectiveness of such initiatives.

The critical quality assessment of included articles has been done by following the criteria stated in the mixed-method appraisal tool (MMAT), that is designed for the appraisal stage of systematic mixed studies reviews. Indeed, it allows to appraise the methodological quality of all the articles included in this review, which include (a) qualitative research, (b) randomized controlled trials, (c) non-randomized studies, (d) quantitative descriptive studies, and (e) mixed methods studies. For each included study, the correct category must be selected for appraisal, followed by an evaluation based on the five criteria specific to that category. For more details on the specific biases investigated for each category, we refer the reader to^17^. The detailed outcomes of this quality appraisal are contained in Table 1, Table 2, Table 3, Table 4, and Table 5, and their statistics in Figure 3.

**Figure 3.**
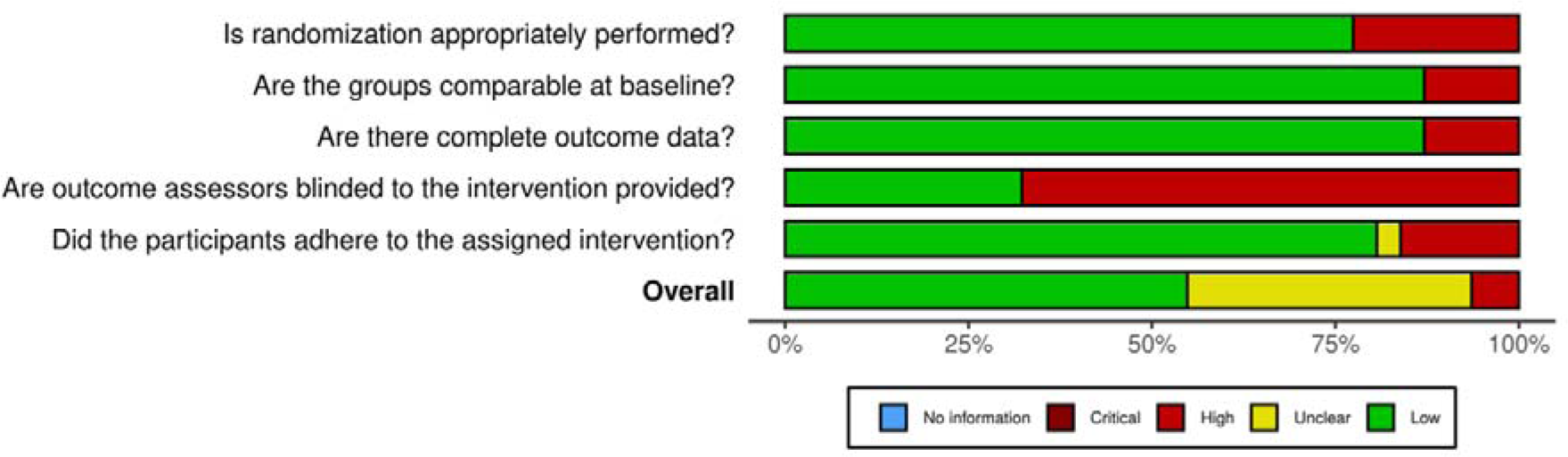
Summary plot of quality assessment for quantitative randomized controlled trials

**Figure 4.**
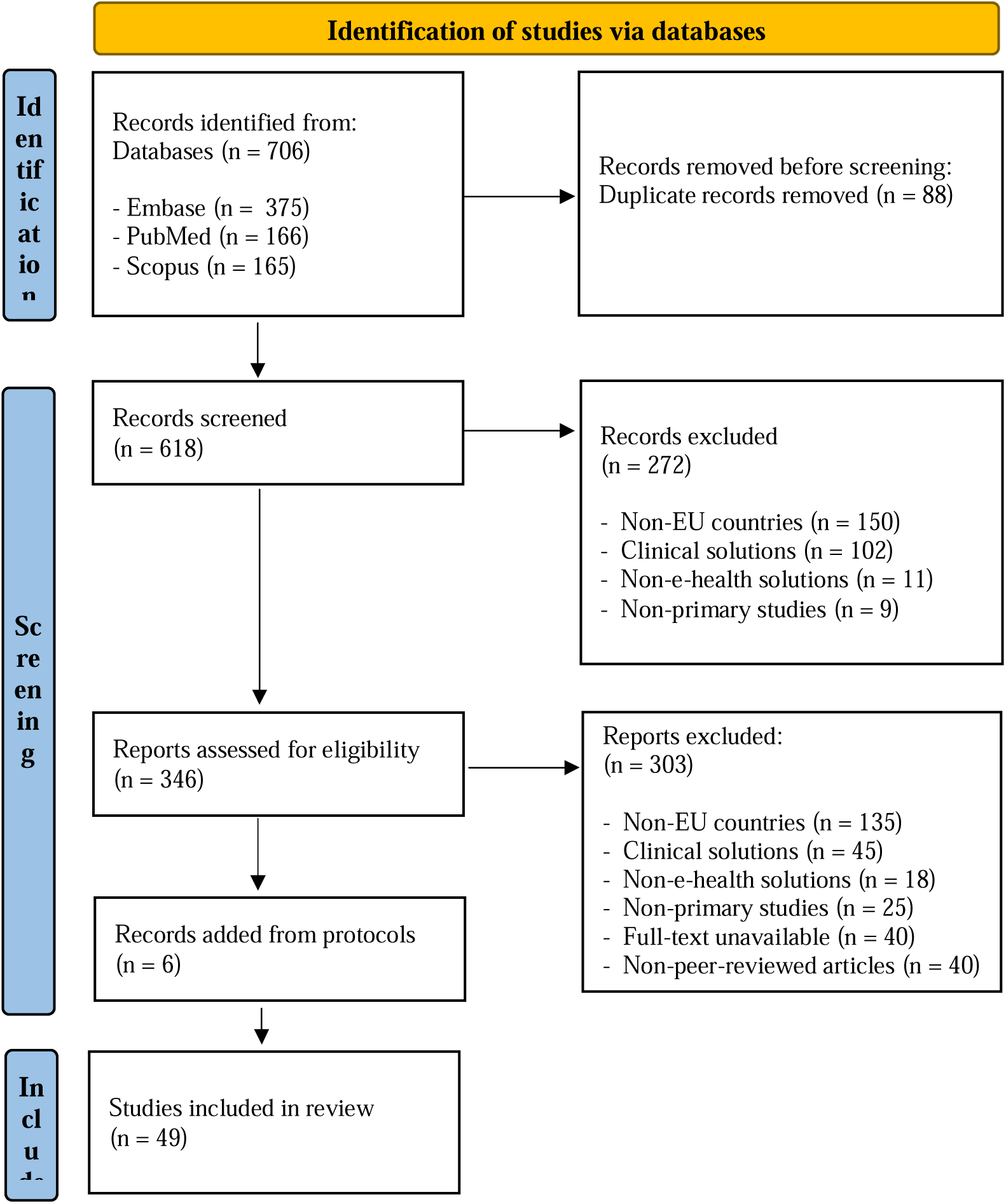
PRISMA flowchart of studies selection.

**Table 1.**
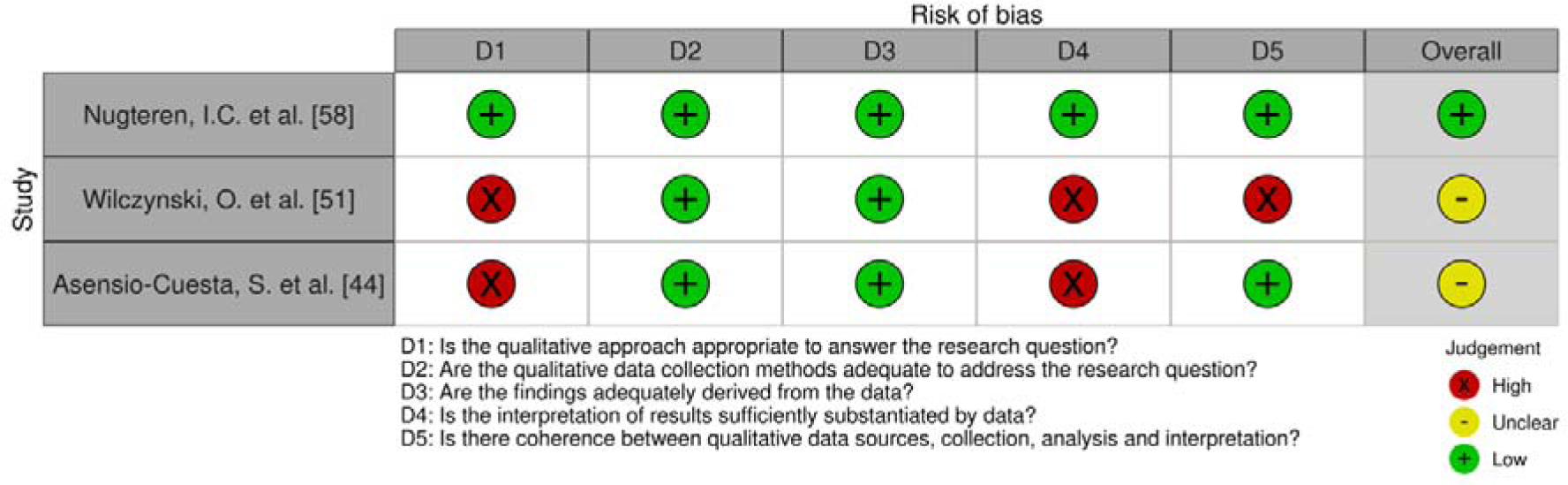
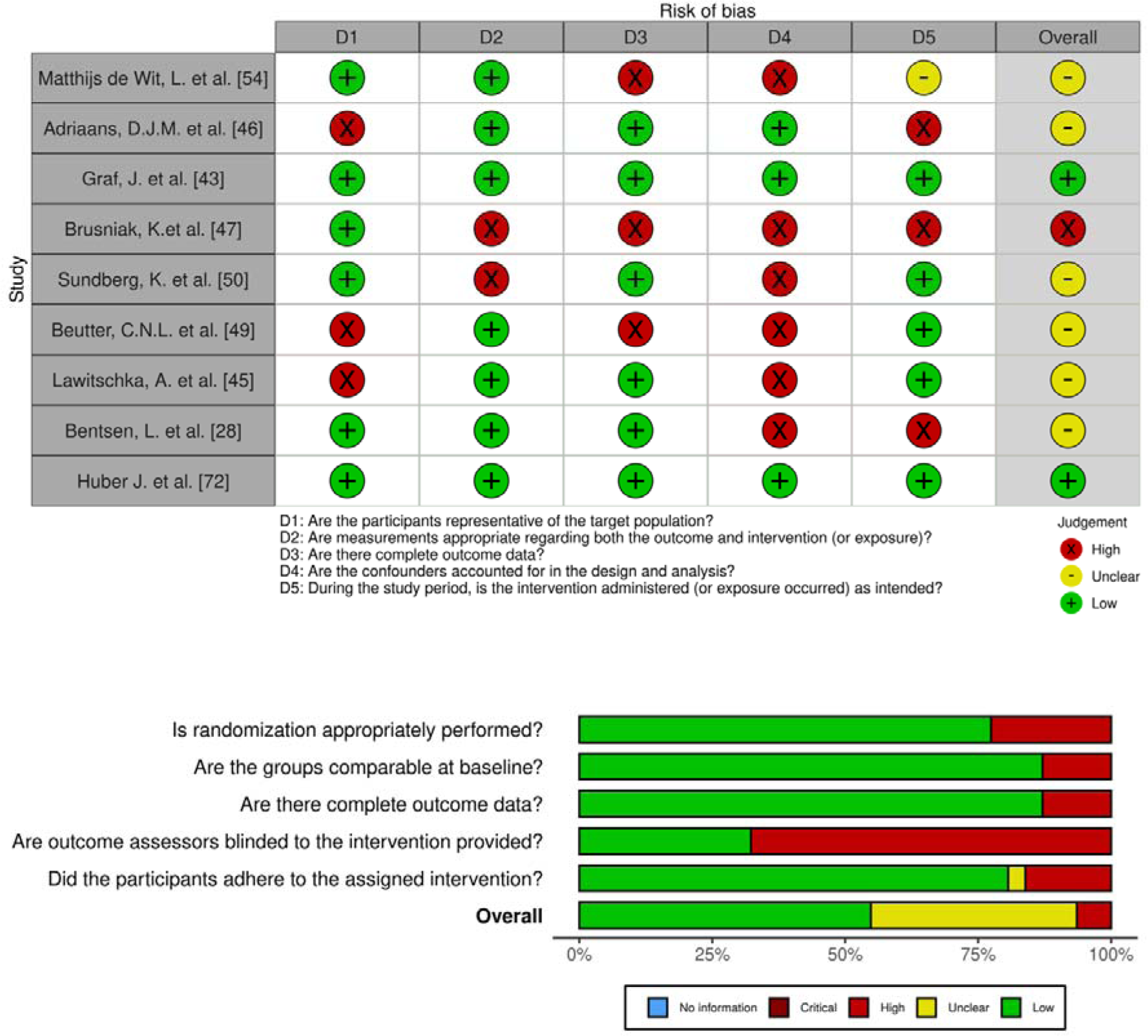
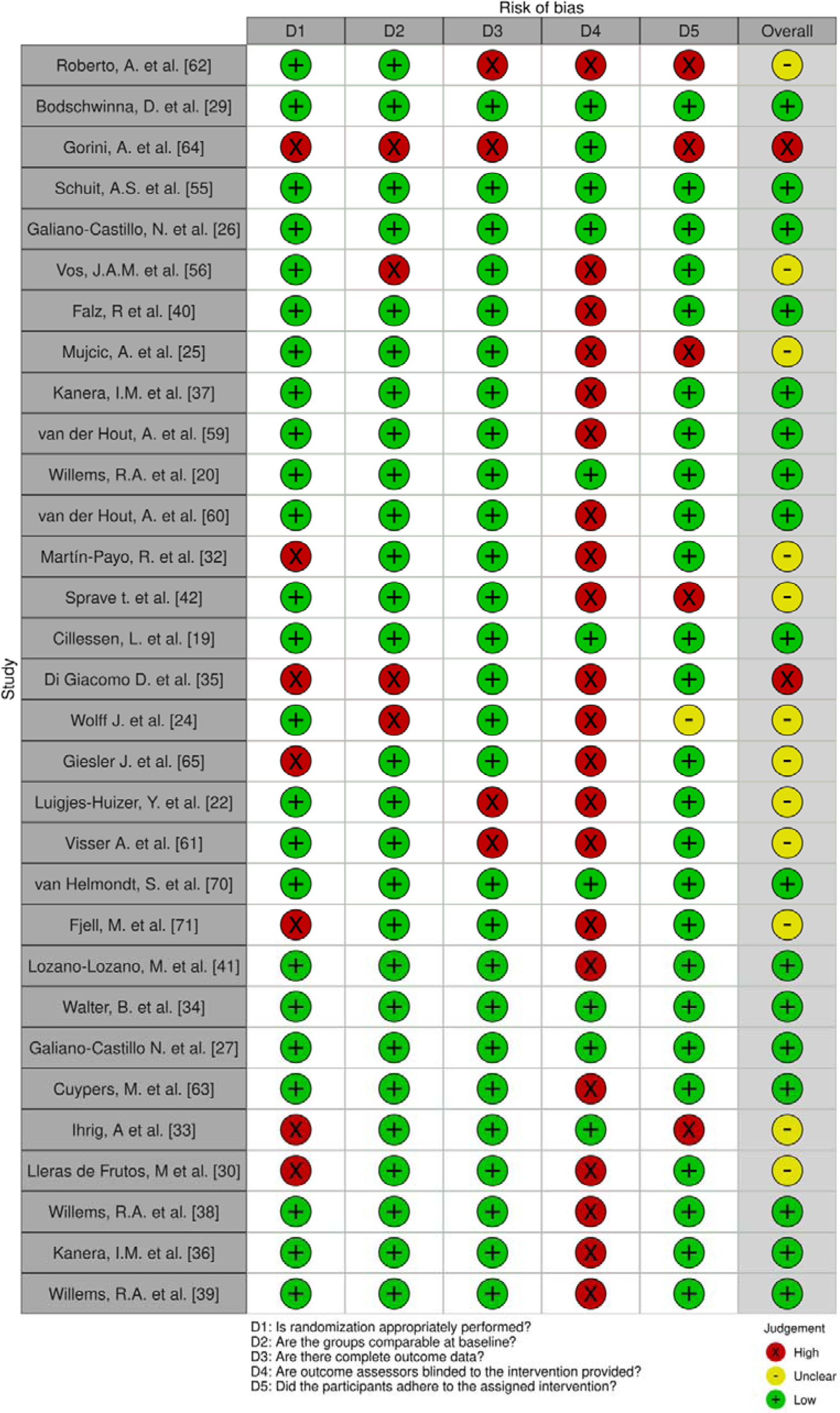
Traffic light table for risk of biases appraisal of quantitative randomized controlled trials.

**Table 2.**
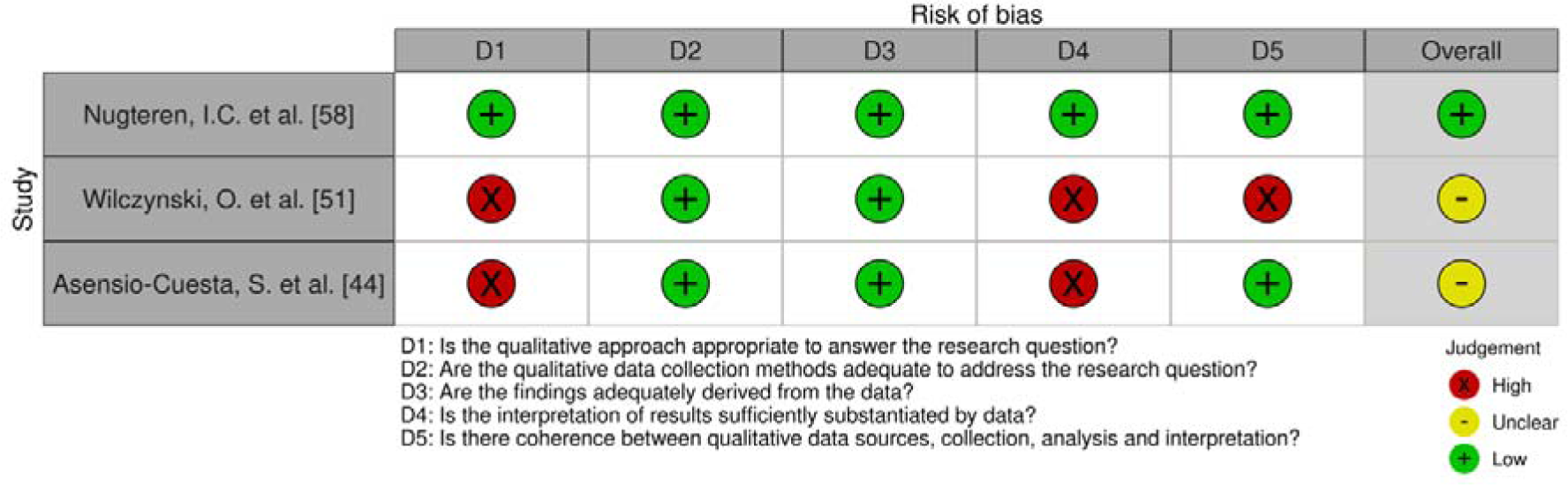
Traffic light table for risk of biases appraisal for qualitative studies.

**Table 3.**
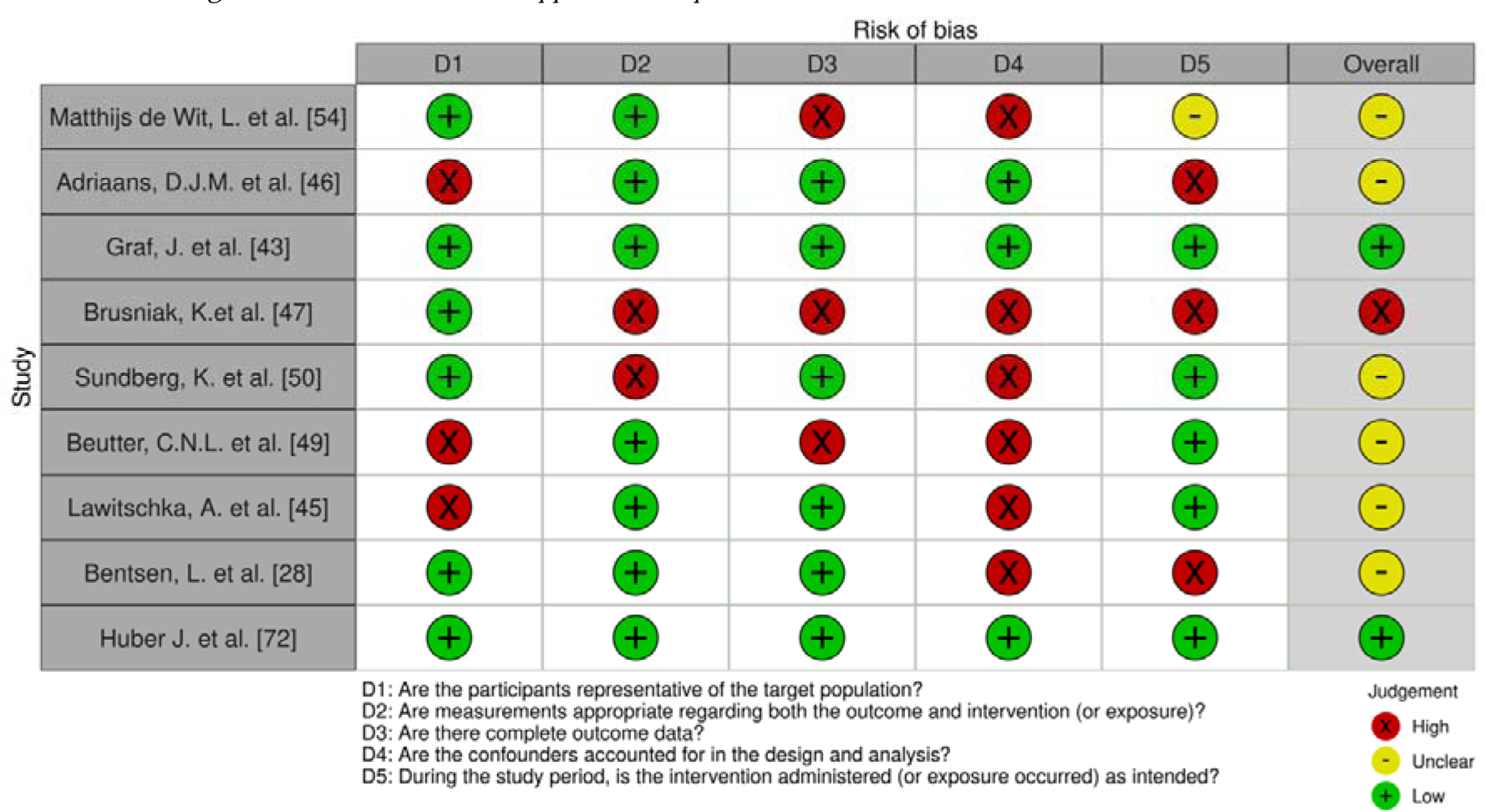
Traffic light table for risk of biases appraisal of quantitative non-randomized controlled trials.

**Table 4.**
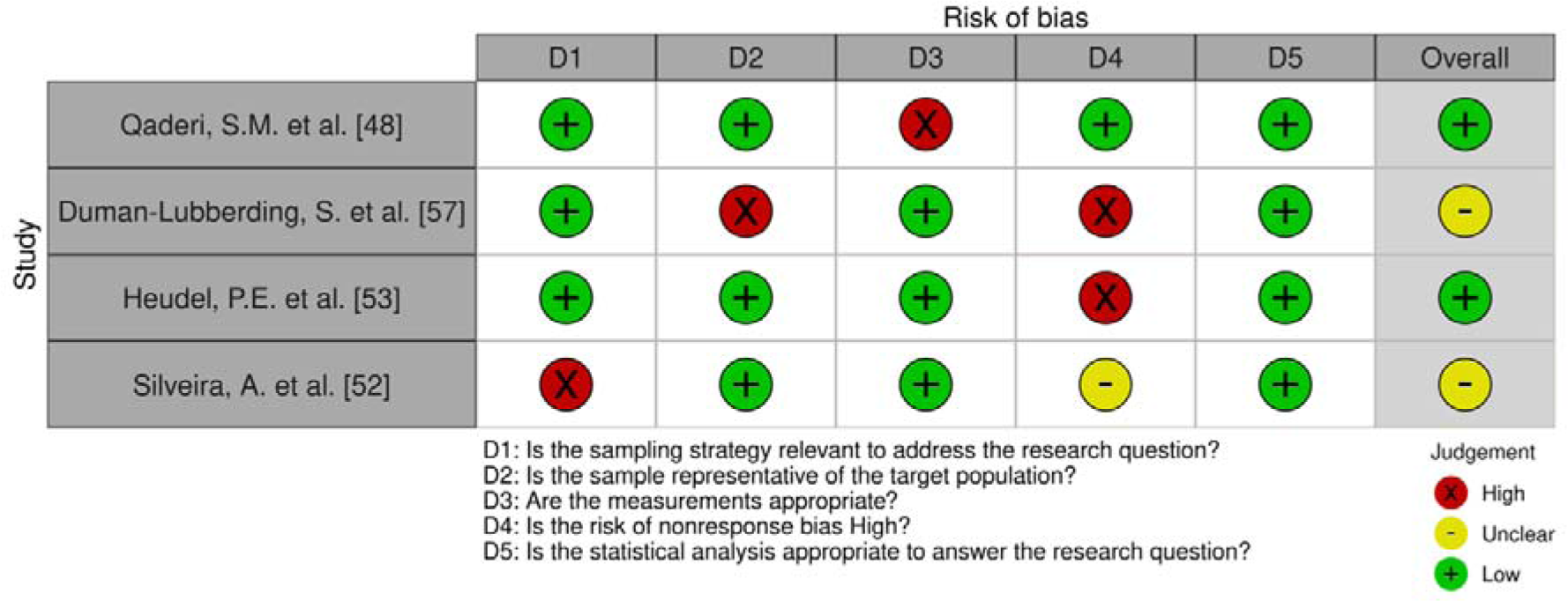
Traffic light table for risk of biases appraisal of quantitative descriptive.

**Table 5.**
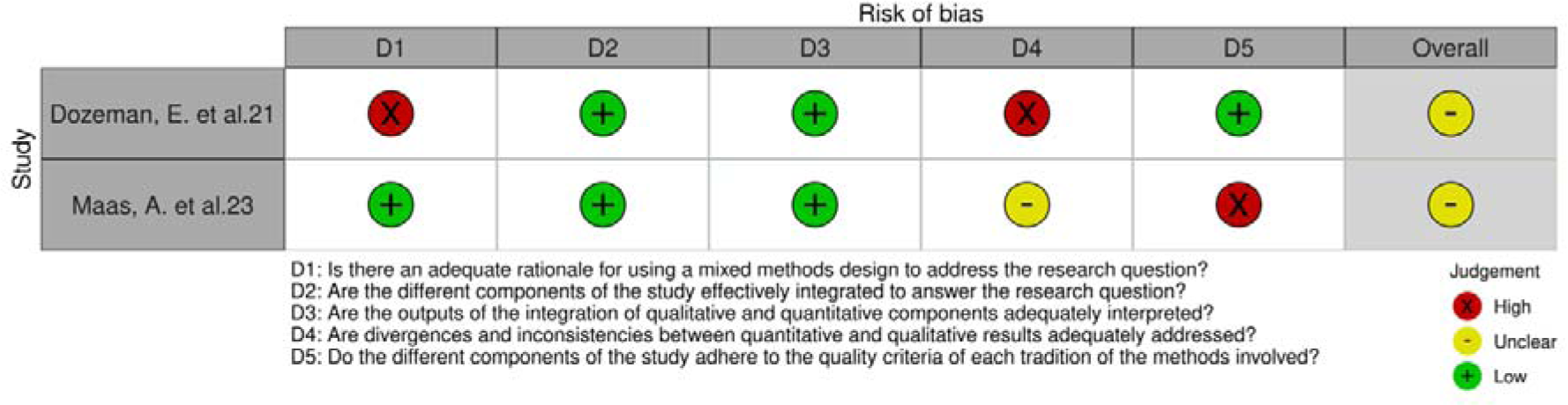
Traffic light table for risk of biases appraisal of mixed-methods studies.

### 2.5 Outline of data synthesis

For all the included studies, their characteristics, the type of digital interventions, and the primary outcomes and results are summarized in Table 6, and then thoroughly described in Section 3, where we provide the answers to each of the three research questions. Namely, after an overview of the study selection in subsection 3.1 the RQ1) on the homogeneity of the distribution of scientific publications across EU countries and cancer types is discussed in subsection 3.2. Then, regarding RQ2), the main areas of digital solutions that emerged from the data extraction have been clustered in (a) psychophysical well-being, (b) reduction of physical cancer treatments side effect, (c) remote monitoring, (d) empowerment and self-efficacy. For each category, we provided a descriptive overview of the findings in subsection 3.3 and discussed RQ3) on the effectiveness of the e-health solutions in subsection 3.4.

**Table 6.** Main characteri stics of the included studies.

| Author and date | Country | Aim of intervention | Study design | Sample | Type of intervention | Feasibility/usability/adherence | Key outcomes | Results |
| --- | --- | --- | --- | --- | --- | --- | --- | --- |
| Roberto A. et al (2020)<sup>62</sup> | Italy | Develop and evaluate a web-based dynamic DA comparing it to with a static brochure (SB). | Pragmatic randomized trial; 2-weeks pretest-posttest; intervention vs control group | Italian woman aged 45-55 who had participated in a screening program in the previous 6 months not having a positive result (N = 2119) | E-health; web-based decision aid | N/A; 20% of participation rate among eligible participants; 47% of adherence rate in follow-up | Informed choice measured on knowledge, attitudes, intentions, participation rate, satisfaction, decisional conflict, and acceptability of decision aid. | DA increased informed choice. DA group had lower decisional conflict compared to the SB group. DA provided more awareness about overdiagnosis compared to SB. DA women had a worse idea of BC mortality frequency. |
| Bodschwinna, D. et al (2022)<sup>29</sup> | Germany | The PartnerCARE online intervention address the support needs of partners of cancer patients | Randomized controlled trial with a two-arm parallel design 4 months follow up ; intervention vs control group | German speaking partners of patients with various cancers (N = 60) | E-health; web-based caregiver guide | 73.3% completion rate. Low study dropout rates at T1 (17%) and T2 (29%). | Feasibility, acceptability measured by dropout rates and individual e-coach feedback; efficacy on psychological distress and anxiety within the intervention group. | High satisfaction with PartnerCARE intervention. Identified positive effects on psychological distress and anxiety within the intervention group. |
| Gorini A. et al (2016)<sup>64</sup> | Italy | Evaluate the effect of an interactive empowerment tool (IEm) on enhancing the breast cancer patient-physician experience, by providing physicians with a personalized patients profile. | randomized controlled trial; intervention vs control group | Adult breast cancer patients who fill in the ALGA-BC (N = 100) | E-health; interactive empowerment tool (IEm) | N/A | Empowerment assessed through a multilingual self-reported questionnaire based on patient knowledge, control, and participation. | Development of a web-based tool for personalized patient profiles. Improvement in patient-physician communication and empowerment. |

**Table 7.**
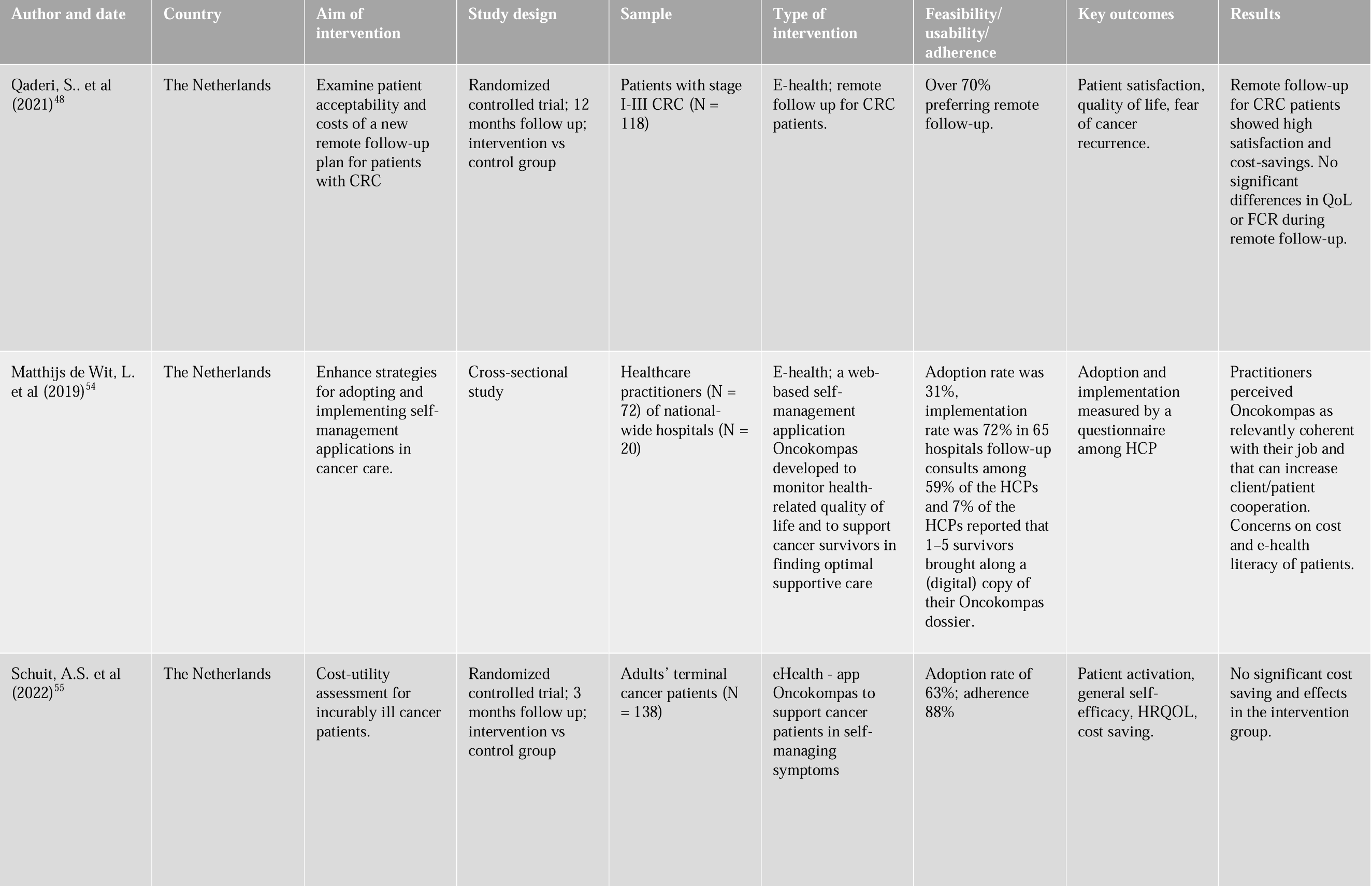
Main characteri stics of the included studies.

**Table 8.**
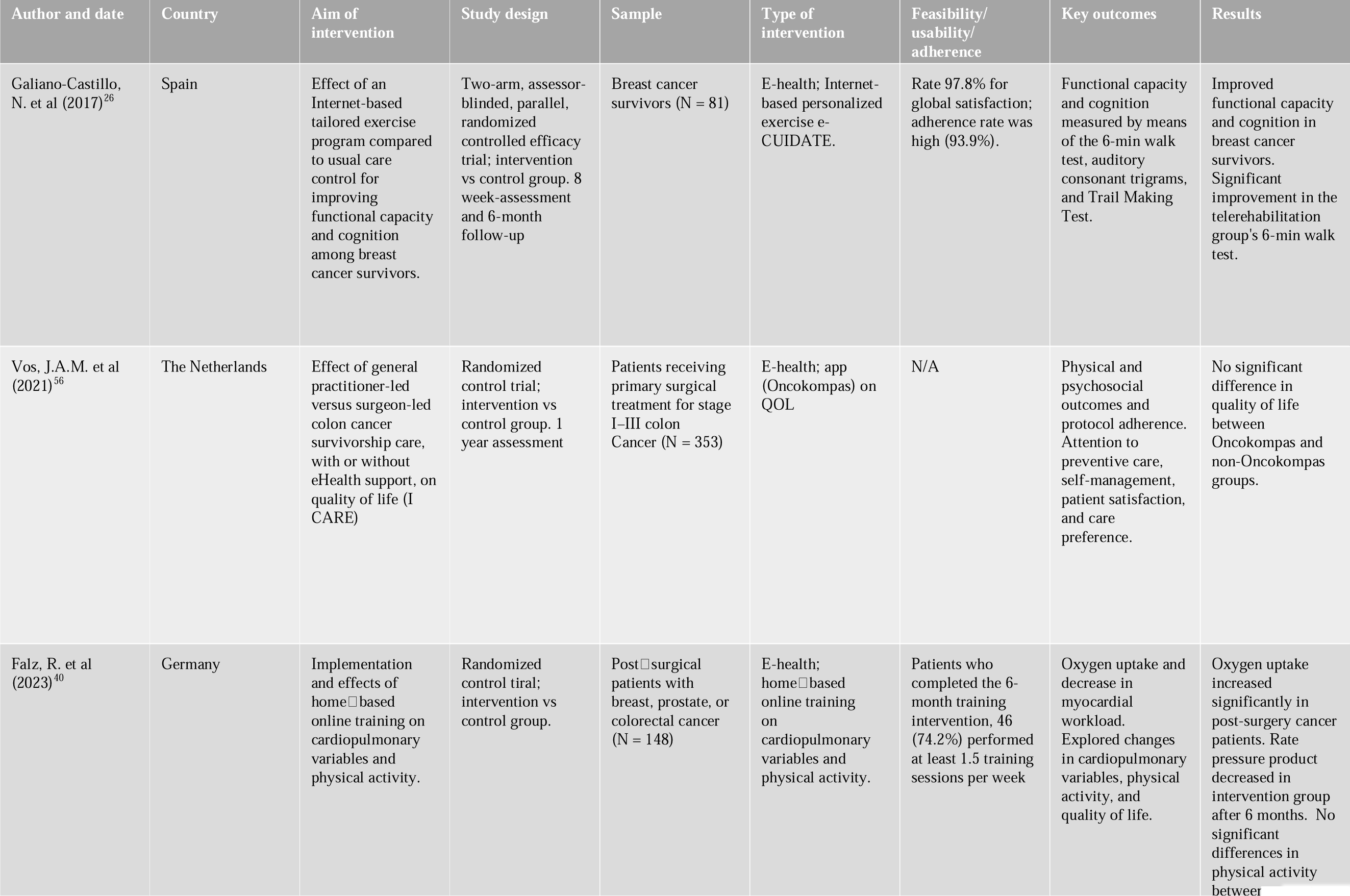
Main characteri stics of the included studies.

**Table 9.** Main characteri stics of the included studies.

| Author and date | Country | Aim of intervention | Study design | Sample | Type of intervention | Feasibility/usability/adherence | Key outcomes | Results |
| --- | --- | --- | --- | --- | --- | --- | --- | --- |
| Mujcic, A. et al (2022)<sup>25</sup> | The Netherlands | Evaluate the effectiveness, cost-effectiveness, and cost-utility of a digital interactive SC intervention compared with a noninteractive web-based information brochure for cancer survivors | Pragmatic 2-arm parallel-group randomized controlled trial | Dutch adult smoking cancer survivors with the intention to quit smoking (N = 165) | E-health; web-based brochure. | 18% eligibility rate; 35% participation rate; 67% adherence rate. | 7-day smoking abstinence at the 6-month follow-up | Quit rates at 6 months were 28% in MyCourse group. MyCourse participants had significantly larger reductions in smoked cigarettes. |
| Duman-Lubberding, S. et al (2016)<sup>57</sup> | The Netherlands | Investigated feasibility of OncoKompas for cancer survivors' self-management. | Pretest-posttest design with survey and interview by a nurse | Adult patients affected by various type of cancer (N = 68) | E-health; digital tool that monitor QoL participant reported outcomes (PROs), generate tailored feedback, and personalize advice on supportive care services. | 64% adoption rate among cancer survivors. 91% of participants used OncoKompas as intended. | Quality of life questionnaire<br>Usage and satisfaction levels | Participants found OncoKompas user-friendly and satisfactory. |
| Dozeman, E. et al (2017)<sup>21</sup> | The Netherlands | Investigate whether guided cognitive behavioral therapy via the Internet (I-CBT) is a feasible and effective solution for this undertreated condition in BRC patients, and to investigate who benefits most. | Open mixed methods design with pre-and post-treatment questionnaires. | Breast cancer patients (N = 100) | E-health; Guided web-based CBT for insomnia in breast cancer patients. | Response rate 56%; adherence rate of 59% | Insomnia severity, fatigue, daily functioning, anxiety, depression were measured. | I-CBT intervention is feasible, accepted, effective for BRC patients. Large to small pre-post effect sizes found on insomnia severity. Younger patients and those with severe insomnia benefited most. |

**Table 10.**
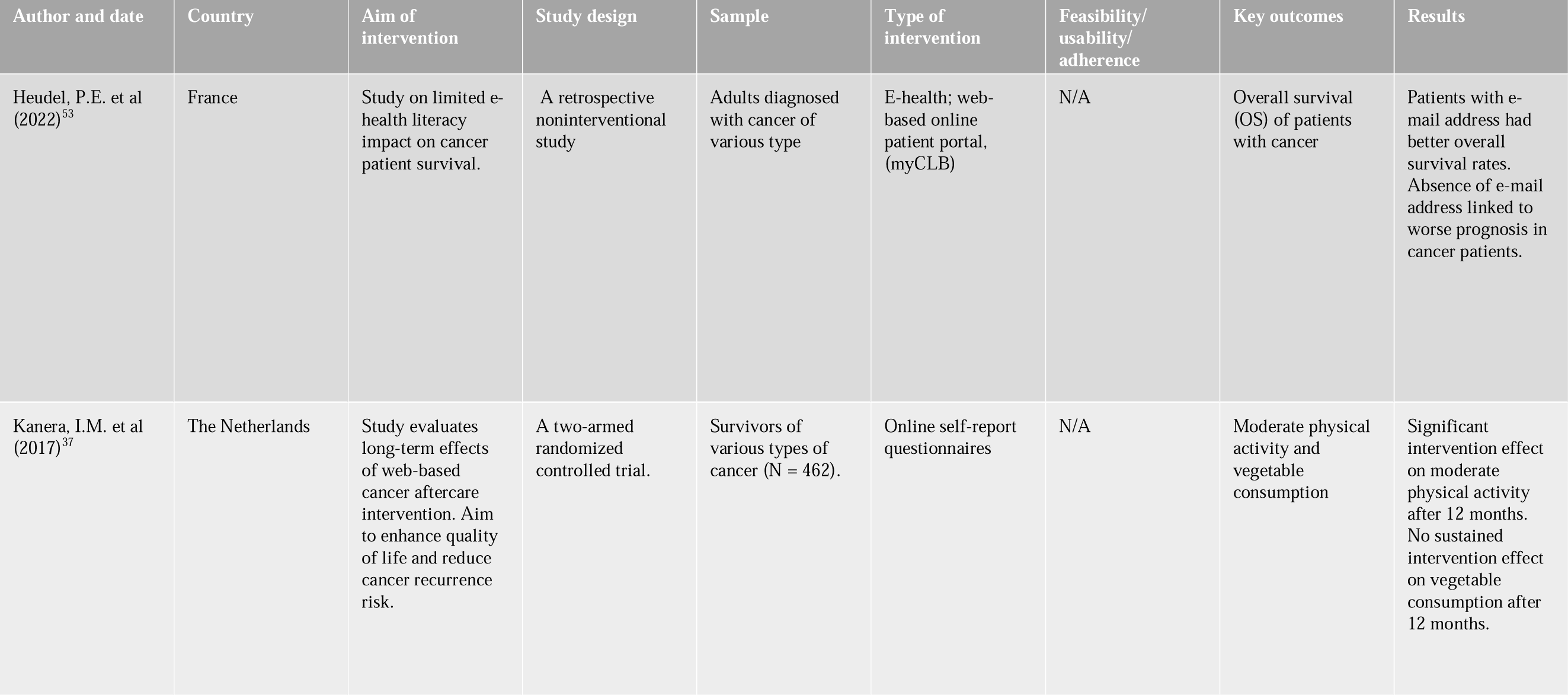
Main characteri stics of the included studies.

**Table 11.** Main characteri stics of the included studies.

| Author and date | Country | Aim of intervention | Study design | Sample | Type of intervention | Feasibility/usability/adherence | Key outcomes | Results |
| --- | --- | --- | --- | --- | --- | --- | --- | --- |
| Nugteren, I.C. et al (2017)<sup>58</sup> | The Netherlands | To investigate patients' opinions about GP involvement in survivorship care and the use of eHealth applications to support self-management. | Qualitative study | Colon cancer patients (N = 20) | Semi-structured interviews | N/A | Opinions on GP involvement and eHealth in colon cancer survivorship care. Participants' satisfaction with current survivorship care and unmet needs. | Patients support GP involvement and Oncokompas 2.0 for survivorship care. Participants see benefits in increased GP involvement and eHealth applications. |
| van der Hout, A. et al (2020)<sup>59</sup> | The Netherlands | Assess the efficacy, reach, and usage of Oncokompas | Randomized control trial; 6-months follow-up | Head and neck cancer patients (N = 625) | E-health, web-based eHealth application that supports survivors in self-management | Adherence of 70% in 6 months follow-up | Patient activation, HRQOL, mental adjustment, supportive care needs, self-efficacy, personal control. | No significant difference in patient activation between intervention and control group. Oncokompas improved HRQOL and tumor-specific symptom burden. |

**Table 12.**
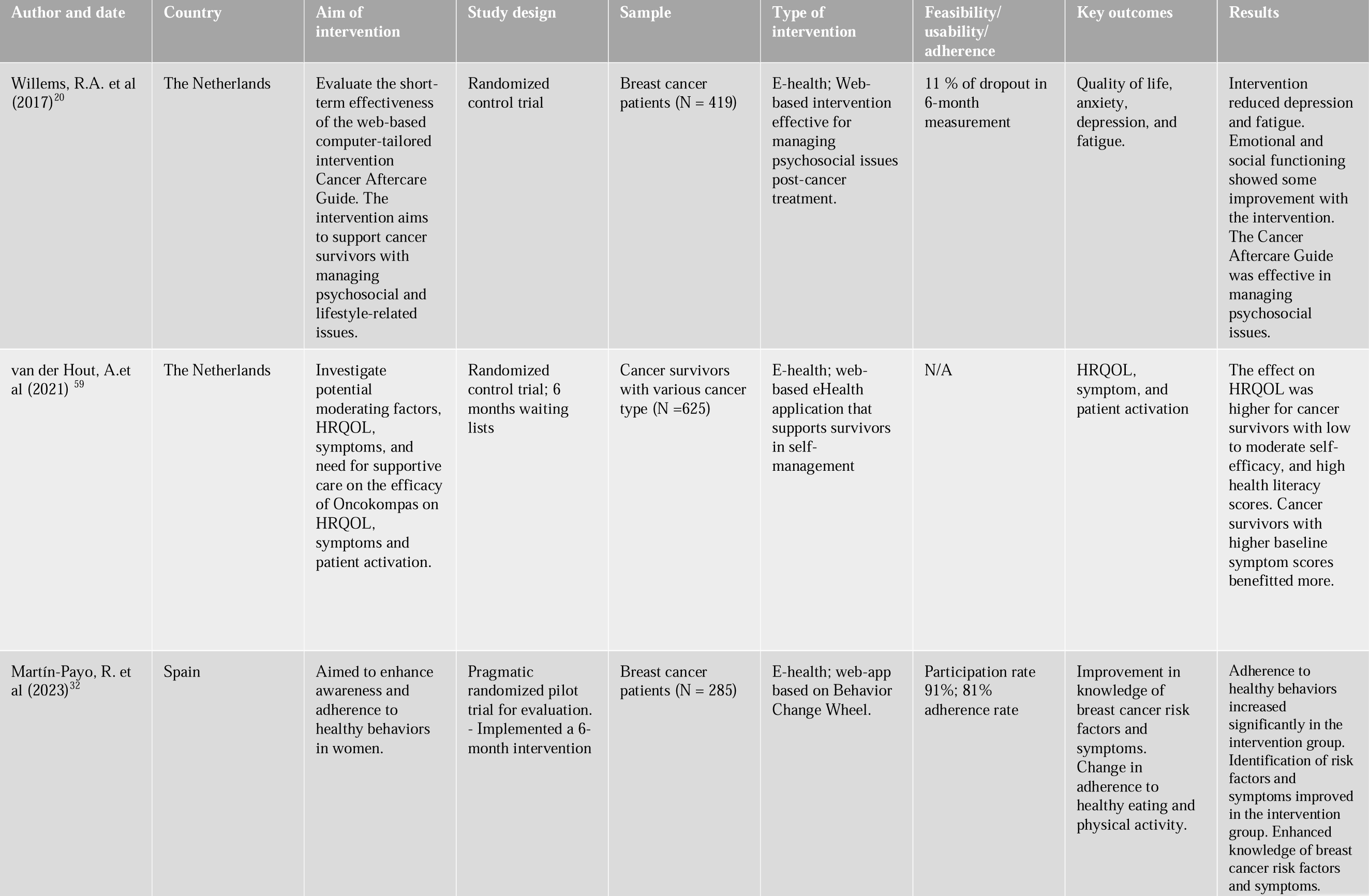
Main characteri stics of the included studies.

**Table 13.** Main characteri stics of the included studies.

| Author and date | Country | Aim of intervention | Study design | Sample | Type of intervention | Feasibility/usability/adherence | Key outcomes | Results |
| --- | --- | --- | --- | --- | --- | --- | --- | --- |
| Sprave, T. et al (2023)<sup>42</sup> | Germany | Investigate the feasibility of integrating electronic patient-reported outcome (ePRO) in the treatment surveillance pathway of HNC patients during radiotherapy. | Randomized control trial between ePRO monitoring or standard-of-care. | Head and neck cancer patients (N = 100) | mHealth; App-controlled monitoring for head and neck cancer patients (APCOT) during radiotherapy. | Feasibility of ePRO monitoring was achieved by all patients in the trial. | Feasibility of ePRO monitoring, patient satisfaction, and symptom burden | Increased head-and-neck cancer-specific symptom burden reported by patients under ePRO surveillance. Improved patient satisfaction in interpersonal manner, financial aspects, and time spent with doctor. |
| Cillessen, L. et al (2018)<sup>19</sup> | The Netherlands | Investigate the long-term effects of MBCT and eMBCT in cancer patients who experience at least mild psychological distress | Randomized control trial; intervention vs control group | Patients with various type of cancer (N = 245) | E-health; online mindfulness-based cognitive therapy for distressed cancer patients. | Adherence rate ranging from 60-70%. | Psychological distress, fear of cancer recurrence, rumination, positive mental health. | Long-term reductions in psychological distress and rumination after interventions. Patients reported less psychological distress after eMBCT compared to MBCT. Patients with low mindfulness skills benefited more from eMBCT. |
| Di Giacomo, D. et al (2021)<sup>35</sup> | Italy | Investigated emotional and mental health impact of scalp cooling technology on oncological patients. | Mixed- method; randomized control trial and observational study | Breast cancer patients (N = 44) | M-health; wearable cooling scalp | 100% of participation rate and adherence | Signs of depression, anxiety, body apperception, body image, expectations, satisfaction assessed. | Digital scalp cooling enhances oncological patients' quality of life. Patients focus on avoiding alopecia, neglecting emotional impact of chemotherapy. |

**Table 14.** Main characteri stics of the included studies.

| Author and date | Country | Aim of intervention | Study design | Sample | Type of intervention | Feasibility/usability/adherence | Key outcomes | Results |
| --- | --- | --- | --- | --- | --- | --- | --- | --- |
| Wolff, J. et al (2023)<sup>24</sup> | Germany | Examined the feasibility and preliminary efficacy of a 12-week digital therapeutic intervention that provides cancer patients with holistic and personalized support to manage distress | Randomized waitlist-controlled pilot study with intervention and control groups. | Patients with gynecological cancer (N = 70) | E-health; Mika app | Dropout rate increased from 15.7% (n=11/70) between baseline and week 4, to 37.1% (n=26/70) between weeks 8-12. The average app usage was 121 min from baseline T1 and decreased over time. | dropout rate, intervention adherence, change in depression, fatigue, and health literacy. | Mika app reduced depressive symptoms by 42% and fatigue by 23.1%. Control group showed no significant changes in health literacy. |
| Giesler, J. et al (2017)<sup>65</sup> | Germany | Evaluate the colorectal cancer module of the German DIPEX website regarding self-efficacy for coping with cancer and patient competence. | Web-based randomized controlled trial with two-group between-subjects design. | Colorectal cancer patients (N = 212) | E-health; module of the German DIPEX website | 78% of adherence. | Self-efficacy for coping with cancer and patient competencies were measured. | No intervention effects on self-efficacy or patient competencies were found. |
| Luigjes-Huizer, Y. et al (2023)<sup>22</sup> | The Netherlands | Assess the effectiveness of a guided online primary care intervention for FCR, compared to waiting list | Randomized control trial; intervention vs control group. | Cancer survivors (N = 173) | e-health; 10-week online programme and three to five video calling sessions with a trained mental health worker. | 81% completed at least half of the intervention by T2; 97% completed the main modules | Fear of Cancer Recurrence severity using the FCRI-SF scale; General mental well-being improvement. | Online intervention reduces Fear of Cancer Recurrence severity significantly. Mental well-being improved significantly in the intervention group. Effectiveness remained at 10 months follow-up. |

**Table 15.** Main characteri stics of the included studies.

| Author and date | Country | Aim of intervention | Study design | Sample | Type of intervention | Feasibility/usability/adherence | Key outcomes | Results |
| --- | --- | --- | --- | --- | --- | --- | --- | --- |
| Visser, A. et al (2018)<sup>61</sup> | The Netherlands | Explores group medical consultations and online support for breast cancer follow-up. Compares My-GMC with individual visits to assess distress and empowerment. | Randomized controlled trial; intervention vs control group. | Breast cancer survivors (N = 109) | E-health; My-GMC, a GMC combined with a tablet-based online app, consisting of three online support group sessions (SGS) and additional information. | The participation rate was 35%. | Psychological distress, empowerment, fear of recurrence, quality of life. | No improvements in distress or empowerment with My-GMC intervention. Significantly more patients experienced peer-support in GMCs than online app. Low satisfaction with the tablet-based online app. |
| van Helmondt, S. et al (2020)<sup>70</sup> | The Netherlands | Evaluates the effectiveness of an online-tailored self-help training based on evidence-based cognitive behavioral therapy principles | Randomized controlled trial; intervention vs control group. | Breast cancer survivors (N = 262) | E-health; the intervention "Less fear after cancer" is a tailored and CBT-based online self-help to reduce FCR | Participation rate 44%; adherence rate 70%. | Fear of cancer recurrence (FCR) severity | No significant effect found on FCR reduction compared to standard care. |
| Fjell, M. et al (2020)<sup>71</sup> | Sweden | Evaluate whether the use of the interactive app Interaktor improves patients' levels of symptom burden and HRQoL. | Randomized controlled trial; intervention vs control group. | Breast cancer patients undergoing neoadjuvant chemotherapy (N = 149) | E-health; Interactive app (Interaktor) for early identification and management of symptoms and to facilitate interaction with health-care professionals | Participation rate 75%; adherence 99%. | Primary endpoints, symptom burden and HRQoL | Statistically significant less symptom prevalence in nausea, vomiting, feeling sad, appetite loss and constipation. |
|  |  |  |  |  |  |  |  | Continues |

**Table 16.**
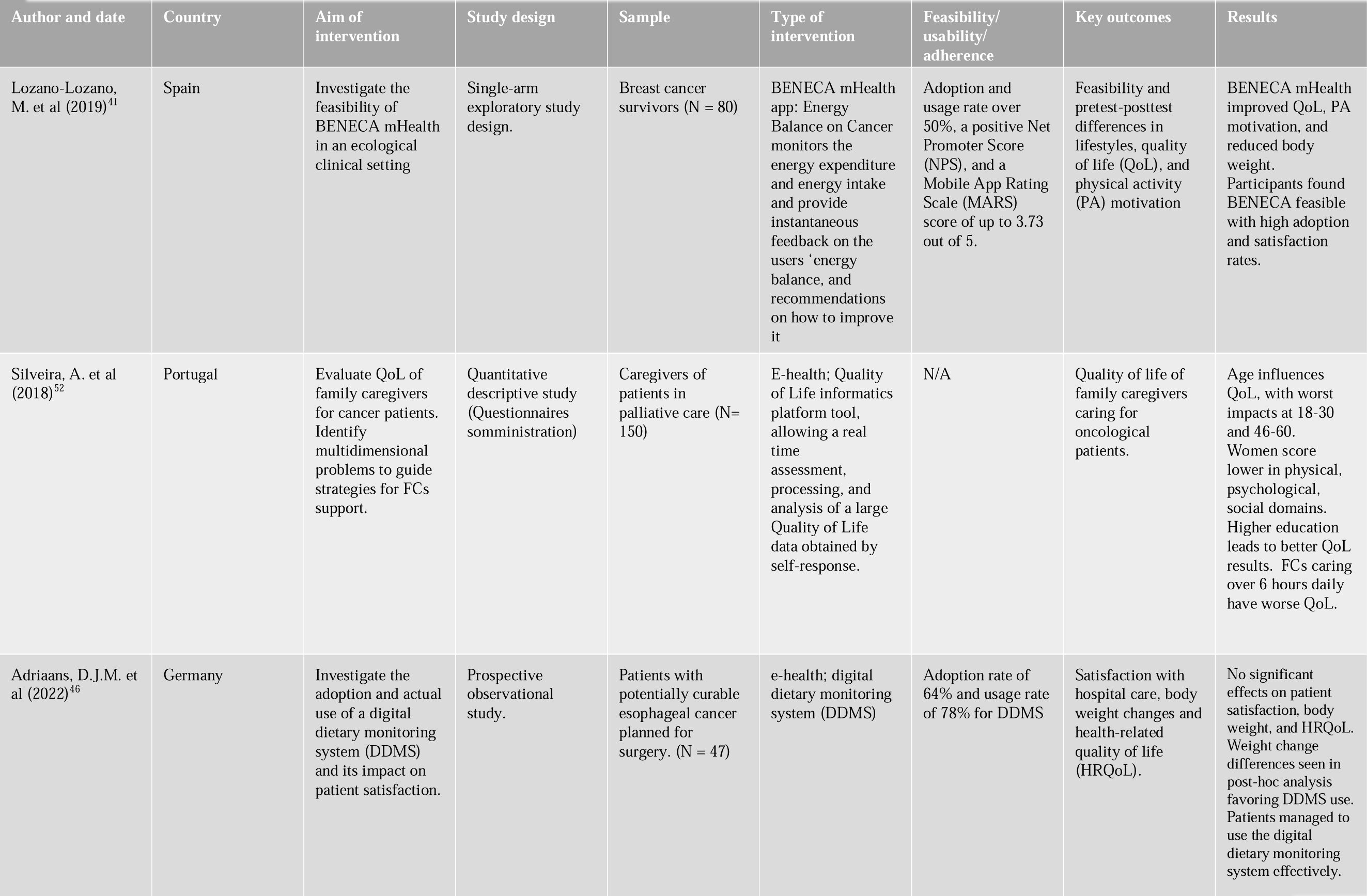
Main characteri stics of the included studies.

**Table 17.** Main characteri stics of the included studies.

| Author and date | Country | Aim of intervention | Study design | Sample | Type of intervention | Feasibility/usability/adherence | Key outcomes | Results |
| --- | --- | --- | --- | --- | --- | --- | --- | --- |
| Graf, J. et al (2022) | Germany | Evaluates the acceptance and evaluation of a tablet-based ePRO app. | 2-center quantitative descriptive study | Women with adjuvant or advanced breast cancer (N = 106) | E-health; electronic patient-reported outcome (ePRO) surveys | N/A | Suitability, stress, and difficulty of ePRO vs. pPRO assessments. Impact of educational level on evaluation of ePROs. | Patients preferred ePRO over pPRO for assessment. Majority found ePRO less stressful and difficult. ePRO improved healthcare in hospitals, according to patients. Educational level influenced the evaluation of ePRO. |
| Wilczynski, O. et al (2022)<sup>51</sup> | France | Describes experiences and expectations of patients treated with ICIs regarding a discussion of HRQoL with health care professionals (HCPs) in cancer management. | Cross-sectional survey | Cancer patients treated with immune checkpoint inhibitors (ICIs), (N = 82) | Online patient community (Carenity) | N/A | Involvement of HCP in a discussion of HRQoL | Patients discussed HRQoL with general practitioners, oncologists, and nurses. Discussions occurred during follow-up, adverse events, and treatment initiation. Patients expected discussions at diagnosis. |

**Table 18.**
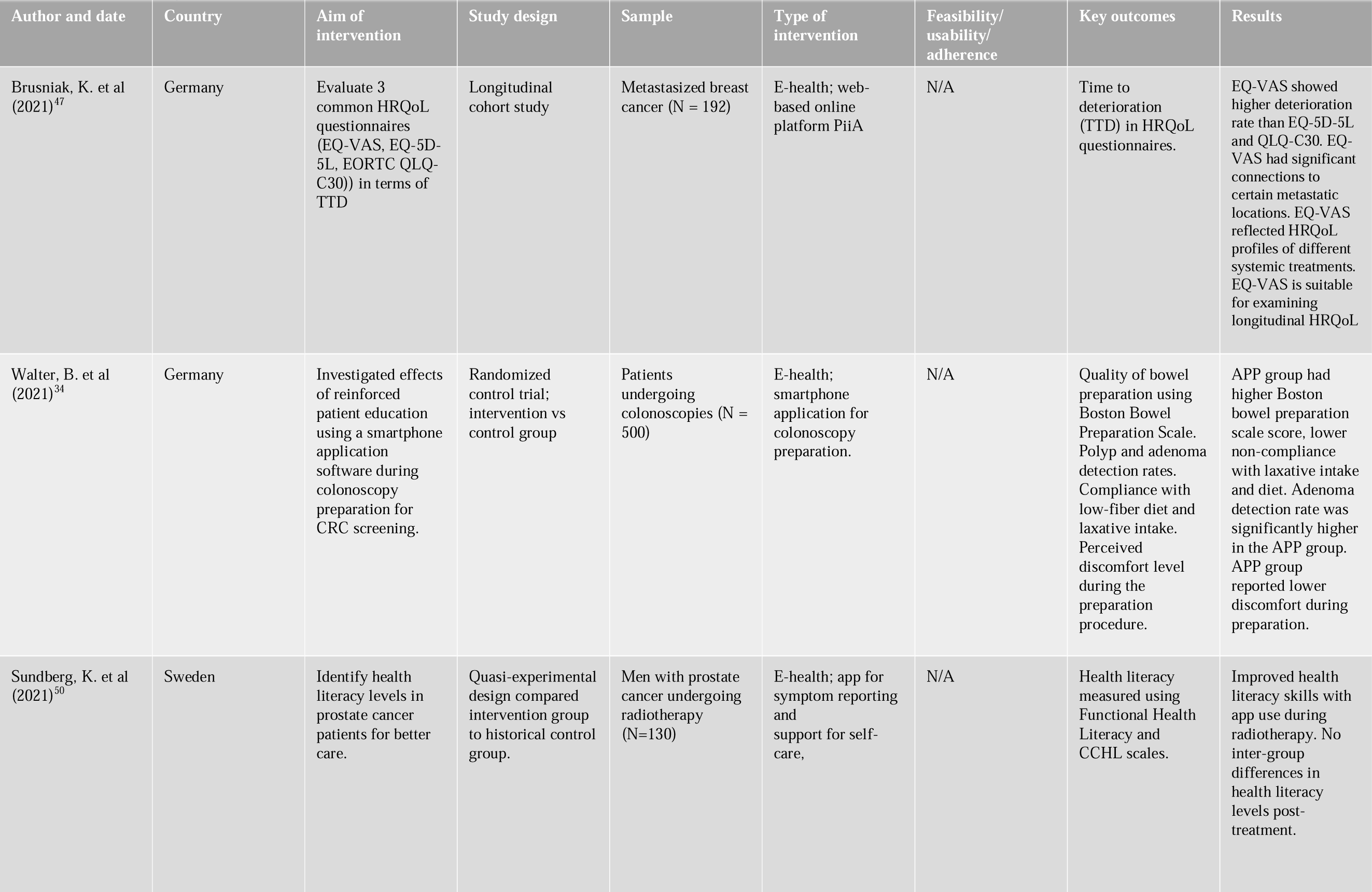
Main characteri stics of the included studies.

**Table 19.**
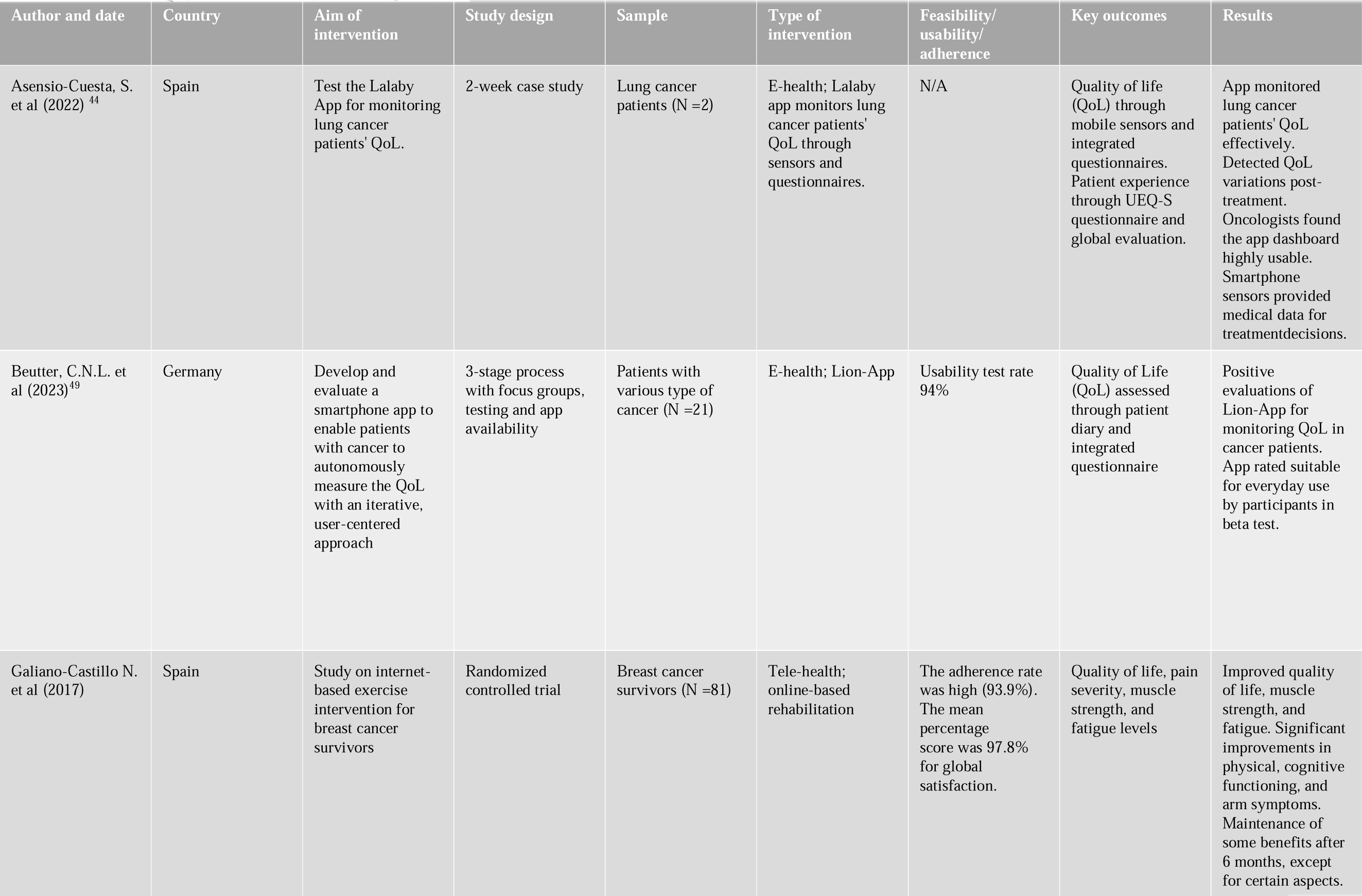
Main characteri stics of the included studies.

**Table 20.**
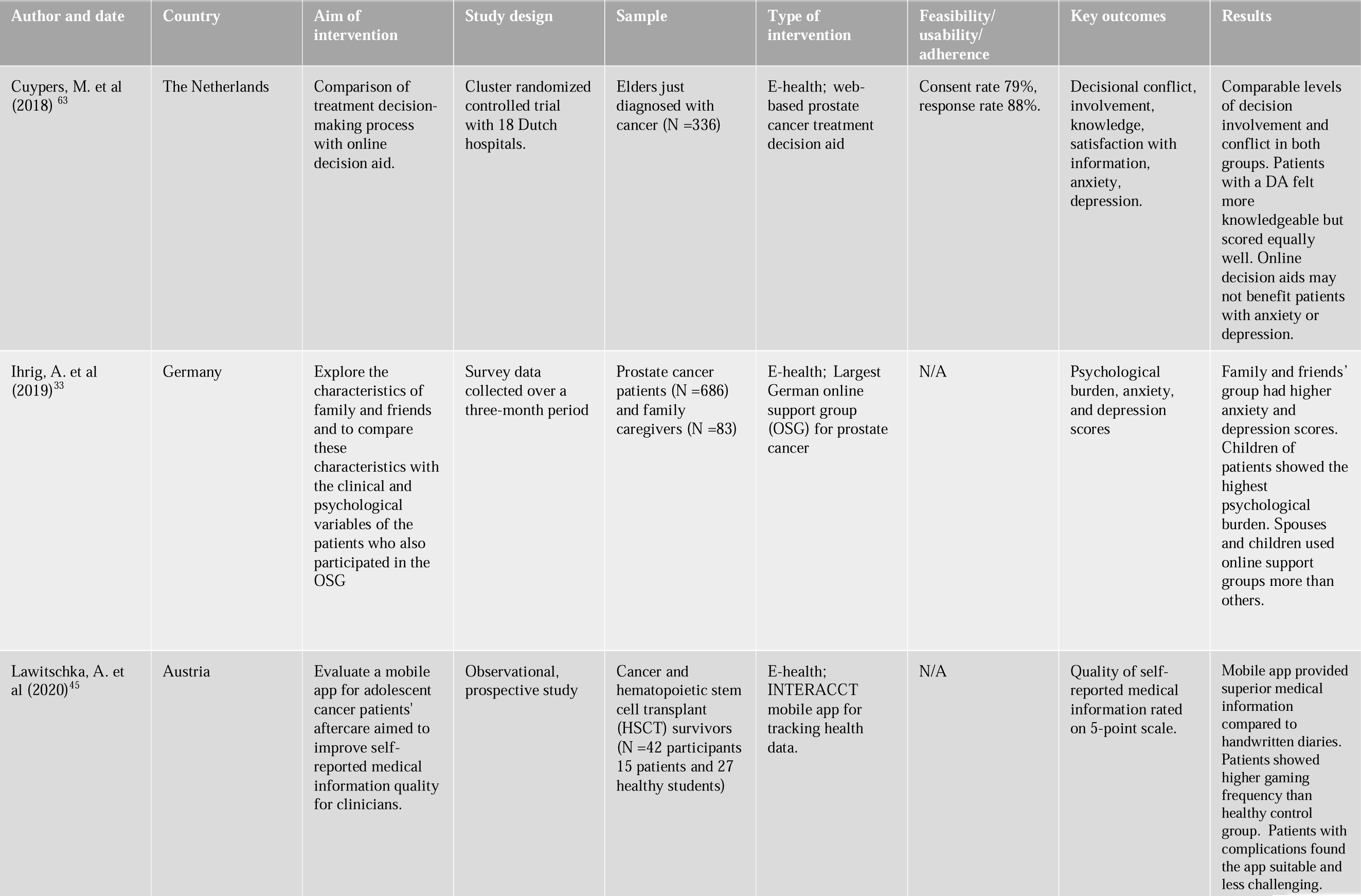
Main characteri stics of the included studies.

**Table 21.**
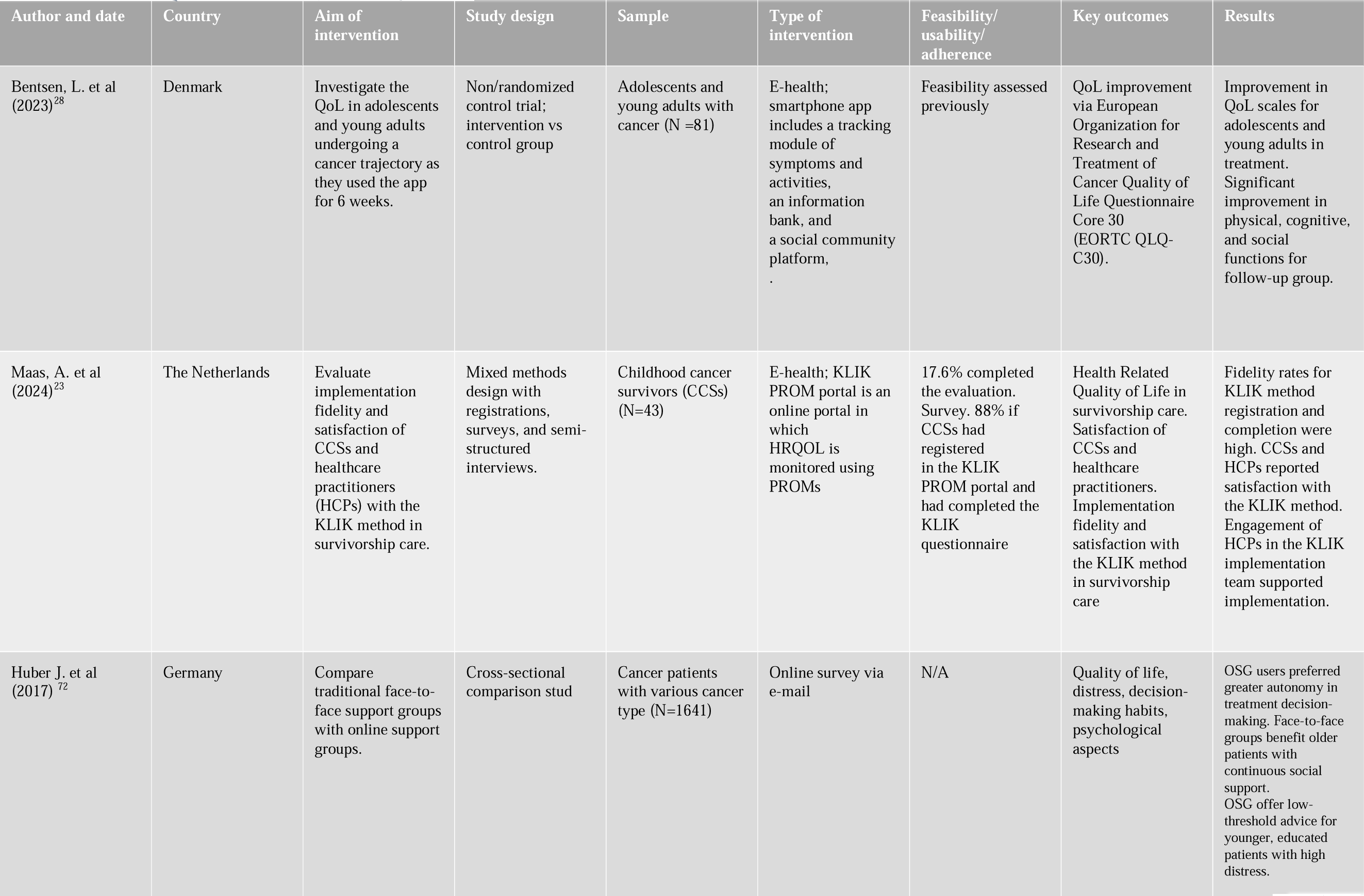
Main characteri stics of the included studies.

**Table 22.**
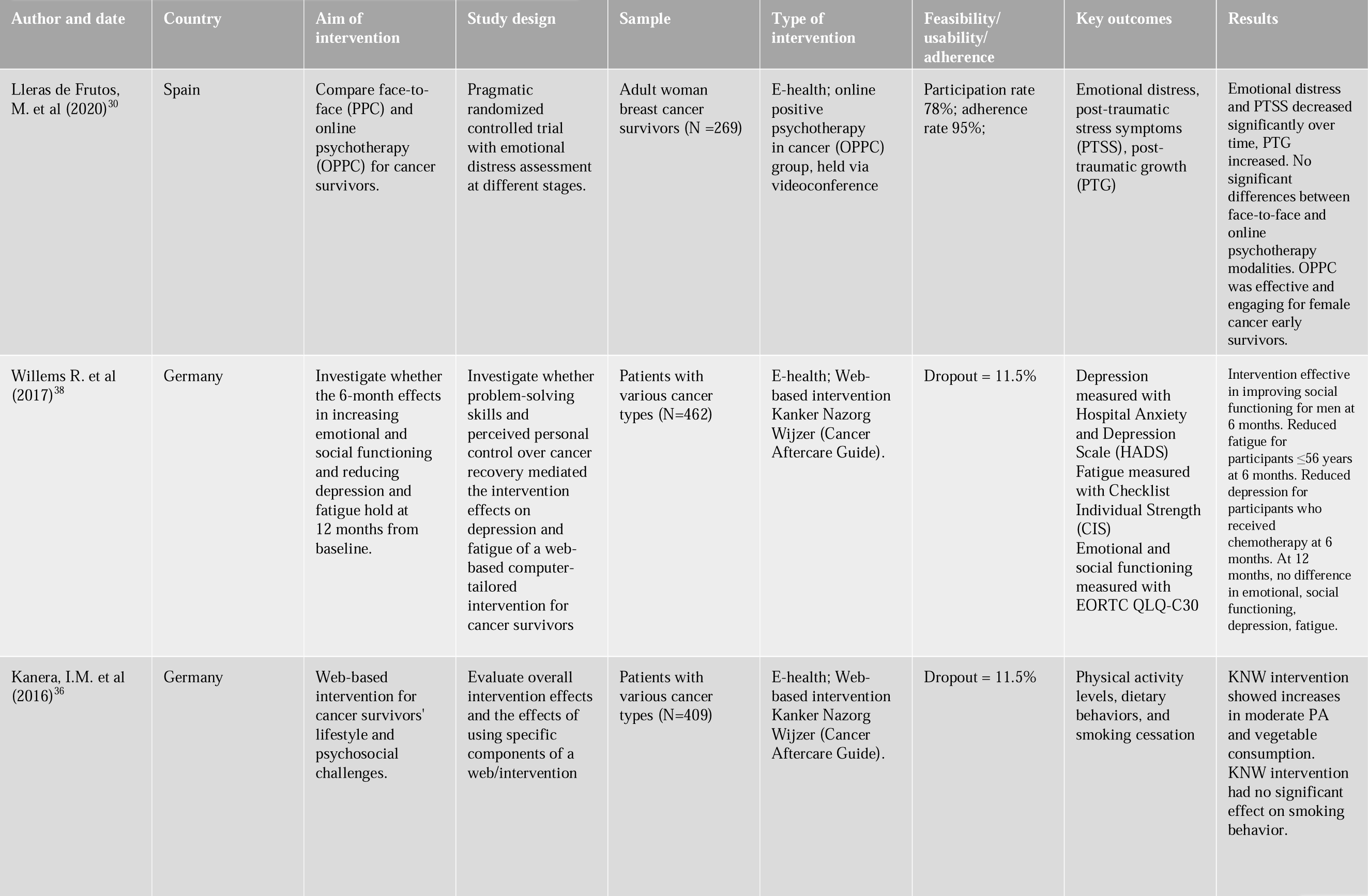
Main characteri stics of the included studies.

**Table 23.** Main characteri stics of the included studies.

| Author and date | Country | Aim of intervention | Study design | Sample | Type of intervention | Feasibility/usability/adherence | Key outcomes | Results |
| --- | --- | --- | --- | --- | --- | --- | --- | --- |
| Willems R. et al (2017)<sup>39</sup> | Germany | Investigate web-based self-management intervention for cancer survivors' effectiveness. | Randomised Controlled Trial with intervention and control groups. | Patients with various cancer types (N=409) | E-health; Web-based intervention | Participation rate 78%; adherence rate 95%; | Depression measured with HADS; fatigue with CIS. | Mediation analyses showed intervention group had lower depression and fatigue. |

## 3. Results

### 3.1. Study selection

Three of the main electronic bibliographic databases, namely Embase, Scopus and PubMed, have been selected to identify studies for the systematic review. The literature search retrieved 706 articles, that after deduplication became 618 for title screening. Following the screening inclusion and exclusion criteria, 276 articles were included in full-text review, and finally 49 have been included in this work.

Note that the reference software manager Zotero has been used to collect the articles included in each stage of the screening process, and in particular the de-duplication has been performed semi-automatically combining the duplicated references identified by the software itself, whereas tags and notes were used to keep track of the exclusion criteria for which some articles have been disregarded. The details of each step are described in Figure.

Across all studies, the number of participants ranged between 2 to 630, with a median of 130 individuals. One strength of this systematic literature review is having a broader viewpoint on the scientific production by encompassing a variety of study types: out of the 49 included studies, the 90% is a quantitative study, 4% of the articles use mixed methods and the remaining 6% is qualitative. Specifically, 31 publications are quantitative randomized control trials, 9 quantitative non-randomized studies (case-studies, cohort studies), 4 quantitative descriptive studies (cross-sectional surveys), 3 qualitative studies (interviews) and 2 mixed-methods studies (quantitative non-randomized trials and qualitative studies).

### 3.2. Answer to RQ1

*The studies proposing digital solutions are more prevalent in the Netherlands and in Germany, and breast cancer patients are the most targeted by single-cancer type interventions*.

The included studies were published between 2014 and 2024, with over 81% of the studies published since 2017 out of a total of 49 articles. Table 6 provides a summary of the included studies. The distribution of academic publications across the EU countries shows a prevalence of studies based in the Netherlands (n = 19; 38%) and in Germany (n = 15; 30%), while the remaining studies were conducted in Spain (n = 6; 12%), Italy (n = 3; 6%), France (n = 2; 4%), Sweden (n = 2; 4%), Portugal (n = 2; 4%), Denmark (n = 1; 2%), and Austria (n = 1; 2%). Moreover, the authors’ nationalities in 96% of the cases corresponded to the country in which the studies have been conducted, highlighting a strong fragmentation and the absence of transnational studies on the impact of digital interventions. Notably, the number of studies across the different countries reflects the rate of cancer incidence (the countries that contributed the most publications on this topic are also the ones most impacted by this illness)^1^.

Nearly half of the studies included were nonspecific with respect to cancer types, encompassing participants with heterogeneous types of cancer (n = 23; 48%), followed by breast cancer (n = 14; 29%), colorectal cancer (n = 5; 10%), head and neck tumor (n = 2; 4%), and the remaining by gynecologic, esophagus, prostate, lung (each corresponding to n = 1; 2%). The high prevalence of breast cancer studies, in alignment with the literature^18^, could be related to the incidence rate and the potential for early detection and intervention, as well as to the high survival rates.

### 3.3. Answer to RQ2

*The key areas of interventions for e-health solutions are i) psychophysical well-being, ii) management of physical distress iii) remote monitoring of vitals and symptoms, and iv) empowerment and self-efficacy*.

The studies included in this review proposed various types of digital intervention for the understanding, support, and enhancement of cancer patients’ and of their caregivers’ needs and quality of life. We clustered these interventions in 4 different thematic areas on the basis of the predominant scope of the digital solutions as follows: a) supporting the psychophysical well-being^19–33^, b) reducing the physical distress induced by the side effects of treatments^20,26,34–41^, c) remote monitoring of vitals and symptoms^42–53^, and d) empowerment and self-efficacy towards patient-centric care^54–65^.

Psychophysical well-being is a crucial aspect to face during cancer care considering the intertwined nature of psychological and physical health. Cancer patients and their caregivers often experience significant emotional stress, including anxiety, depression, and solitude, which can exacerbate their physical symptoms and affect their overall quality of life. E-health solutions, such as mobile applications offering mindfulness exercises, virtual therapy sessions, and online support communities, provide accessible and tailored mental health resources that are available every day and in every moment of the day, and that would be impossible to provide by means of traditional healthcare.

The management of physical distress is another critical area addressed by e-health interventions for oncological patients. Cancer treatments, including chemotherapy, radiation, and surgery, frequently result in side effects such as pain, fatigue, nausea, hair loss, weight gaining and so on. E-health tools, like online exercise applications and dietary management platforms, as well as wearable devices for some specific issues like alopecia, enable patients to cope with physical discomfort and embrace a healthier lifestyle, and more importantly to help them be consistent with it over a longer period.

Remote monitoring of vitals and symptoms is a key component of e-health solutions, offering a proactive approach to patient care. Mobile health apps and web-based interventions allow for the autonomous log of patients’ symptoms in real-time and the timely medical guidance. This real-time data collection enables healthcare providers to monitor patients’ health status remotely and intervene early when abnormalities are detected. The ability to oversee patients’ conditions without requiring frequent hospital visits not only reduces the burden on caregivers and healthcare facilities but also minimizes patient exposure to hospital environments, which is particularly beneficial for immunocompromised individuals.

Enhancing empowerment and self-efficacy can improve the attitude of patients and their caregivers in the difficult moment of cancer treatment and beyond. E-health solutions in this area can include educational resources that provide detailed information about cancer types, treatment options, and self-care strategies, interactive platforms and decision-aid tools that encourage patients to actively participate in their health care by tracking progress, exchange experiences and advice and making informed decisions. This empowerment fosters a sense of control and confidence in the patients, leading to a better adherence to treatment regimens and proactive health behaviors, which are associated with improved health outcomes.

### 3.4. Answer to RQ3

*he effectiveness of digital solutions is typically higher than traditional healthcare, especially those focusing on psychosocial well-being*.

In what follows, we report the results in terms of effectiveness of the digital health solutions identified in this systematic literature review clustered by thematic areas identified in the previous section.

#### 3.4.1. Digital interventions for improving psychophysical well-being

In study^19^, the authors proved the effectiveness of electronic mindfulness behavioral cognitive therapy (eMBCT) in reducing psychological distress with respect to traditional MBCT, especially for patients with low mindfulness skills. Willems *et al.*^20^ proved a positive impact on emotional and social functioning, and showed a decrease in depression and fatigue 6 months after baseline, which remained significant also when considering effect size.

Dozeman *et al.*^21^ proved that a guided web-based cognitive therapy for insomnia (I-CBT) in breast cancer patients is feasible and effective for younger breast cancer patients and those with severe insomnia. Luigjes-Huizer *et al.*^22^ showed how online primary care intervention for reducing fear of cancer recurrence (FCR), consisting of an e-health program and video calling sessions with a mental health worker, improve the general mental well-being and FCR severity, with the effectiveness of the intervention remaining at 10 months follow-up. The outcomes across different studies, however, are mixed, as in another study no effect of CBT-based online self-help training on FCR has been found^23^.

The digital therapeutic MIKA app based on holistic and personalized approach demonstrated efficacy in reducing depression and fatigue by 42% and by 23.1%, respectively, with respect to the control group^24^. The web-based intervention MyCourse-Quit smoking, based on CBT, acceptance and commitment therapy (ACT) and motivational interviewing (MI) techniques, proved to be more effective in reducing the daily consumption of cigarettes with respect to control group in cancer survivors^25^. The internet-based tailored rehabilitation exercise program e-CUIDATE improved functional capacity in terms of distance covered in the 6-min test and cognition in terms of memory in survivors that participated to the intervention^26,27^. The Danish smartphone app, Kræftværket, includes a tracking module for symptoms and activities, an information bank with both text and video material, and a social community platform that facilitates networking and sharing experiences; it showed improvement in QoL scales for young adults in treatment, and in physical, cognitive, and social functions for follow-up group^28^. The comparison of face-to-face and online psychotherapy or support groups for cancer survivors showed no significant difference, thereby suggesting the potential of online interventions to overcome geographical barriers^30,31^. Martín-Payo *et al.*^32^ demonstrated how a web-app based on Behavior Change Wheel Model improved adherence to healthy behaviors, and significantly facilitated the identification of risk factors and symptoms in the intervention group of breast cancer patients.

Two studies also focused on informal caregivers^29,33^: the first German app to address informal caregivers needs has been the PartnerCare app^29^, which showed positive effects on psychological distress and anxiety within the intervention group by means of psychoeducation, behavioral therapy, supportive therapy and guided imagery. A second study focused instead on the German online support group (OSG), which is a forum for counselling, dissemination and treatment between prostate cancer patients and informal caregivers. Namely, Ihrig *et al.*^33^ have found OSG to be beneficial in terms of psychological burden for informal caregivers of cancer patients’, that are the most affected by this type of distress.

#### 3.4.2. Digital interventions for managing physical distress

An app for patient education, improving compliance and discomfort levels for colonoscopy preparation for screening has been introduced in^34^, showing positive effects in all the outcomes measured. The digital scalp cooling technique effectively prevented chemotherapy-induced alopecia, thereby enhancing psychological well-being in cancer patients^35^. The web-based intervention Cancer Aftercare Guide (KNW) is a computer-tailored intervention that aims to increase survivors’ quality of life that comprises 8 separate modules that target the topics physical activity, diet, smoking cessation, return-to-work, fatigue, anxiety and depression, social relationships, and residual problems^20,36–39^. It was shown to be effective in improving social functioning, reducing depression and fatigue at 6 months, and increasing moderate physical activity of cancer survivors younger than 57 years. However, it was not effective in fostering vegetable consumption, smoking cessation, emotional and social functioning, and in reducing depression and fatigue after 12 months.

According to^40^, home-based online training with video presentations for post-surgical cancer patients are effective in enhancing oxygen uptake and decreasing myocardial workload during exercise. Lozano-Lozano *et al.*^41^ developed the app BENECA (Energy Balance on Cancer) to stimulate changes in breast cancer survivors’ lifestyles based on energy balance. Namely, their goal was to monitor the energy expenditure and energy intake of breast cancer survivors and provide instantaneous feedback on the users’ energy; the outcome was the study was an improved QoL, PA motivation, and reduced body weight, with high adoption and satisfaction rates among the participants. Galiano-Castillo *et al.*^26^ also focused on lifestyle in breast cancer patients, showing how an internet-based exercise intervention yielded significant improvements in quality of life, muscle strength, fatigue, physical and cognitive functioning, and arm symptoms. Such an improvement remained after 6 months.

#### 3.4.3. Digital interventions for remote monitoring

Sprave *et al.*^42^ demonstrated the feasibility of integrating app-based electronic patient-reported outcomes (ePRO) in patients with head and neck cancer (HNC) undergoing radiotherapy, increasing reporting of cancer-specific burden and improved patient satisfaction. Graf *et al*.^43^ also reported an improved acceptance and evaluation of a tablet-based ePRO app when compared to paper-based patient reported outcome (pPRO), with patients finding ePRO assessment less stressful and less difficult. Asensio-Cuesta *et al.*^44^ focused on the Lalaby app that monitors quality of life of lung cancer patients through sensors and questionnaires in real time, and found it effective towards better therapeutic decisions. The use of ePRO apps can be however hindered by a limited health literacy: Haudel *et al.*^53^ showed how having an account on an online portal for ePRO (which is a proxy for health literacy) is strongly related to lower overall survival rate, thereby stressing the potential value in the use of ePRO apps. In an effort to enhance health literacy, Sundberg *et al.*^50^ supported the use of an interactive app for prostate cancer symptom management during radiotherapy.

E-health solutions can also be devised to enhance the quality of the information provided by adolescent cancer patients to clinicians: Lawitschka *et al.*^45^ showed how the web-based gamified mobile app INTERACCT (Integrating Entertainment and Reaction Assessment into Child Cancer Therapy) enables adolescent cancer patients to self-track in real-time symptoms improving quality of medical information for clinicians compared to traditional methods.

Digital solutions have also been implemented to monitor health-related quality of life (HRQoL). Adriaans *et al.*^46^ tested an online platform, the KLIK portal, can be used to monitor HRQoL by using patient reported outcome measures (PROMs). Namely, they tested the digital dietary monitoring system for esophageal cancer patients, without showing any significant effects on patient satisfaction, body weight, and HRQoL. Brusniak *et al.*^47^ evaluated HRQoL in metastatic breast cancer patients using digital monitoring, allowing for the inclusion of patients not living in close proximity to the care center, not only in conjunction with treatment. HRQoL was measured through the administration of 3 commonly used questionnaires. Beutter *et al.*^49^ developed and tested a smartphone app called the Lion-App, through which patients with various type of cancer can autonomously measure the QoL with an iterative, user-centered approach, suitable for a daily use. Wilczynski et al.^51^ focused on the Carenity online patient community in which patients and caregivers can share their experiences, exchange information and advice and also participate in online surveys concerning various aspects of disease perceptions. This platform allowed patients’ to discuss HRQoL with practitioners starting from diagnosis, and not only when side effects arise. Silveira *et al.*^52^ focused instead on QoL monitoring for caregivers of oncological patients admitted to the Palliative Care Service of the Portuguese Oncology Institute of Porto. They used a Platform for QoL assessment in oncology, named OpQoL, showing that the worst parameters in terms of QoL were observed for female caregivers between 18-30 and 46-60 years. The scores worsen when one gives care for more than 6 hours a day, whereas higher education is associated to better QoL results.

Remote applications are also an opportunity of reducing the costs while maintaining the same quality of care. Qaderi *et al.*^48^ showed that remote follow-up can yield significant cost-savings without compromising quality of care for colorectal cancer patients.

#### 3.4.4. Digital interventions for empowerment

A very well-studied web-based application towards patients’ empowerment is the Dutch app Onkocompas^54–59^. Specifically, Onkocompas is a web-based self-management application where participants reported outcomes are used to then provide by personalized feedback and self-care advice to stimulate patient activation. The feasibility, acceptability, short- and long-term effectiveness and cost reduction associated to the use of the application have been thoroughly analyzed across several studies^54–59^. Namely, Nugteren *et al.*^58^ showed that the patients’ express a positive attitude towards the use of Onkocompas, together with the need for a greater involvement of general practitioners in survivorship care. In^57^ the feasibility of this app for cancer survivors’ self-management has been assessed with 64% adoption rate among cancer survivors, showing high satisfaction and usability; then, in^54^, the app has been tested for 1-year in a national pilot in The Netherlands, with a nationwide adoption rate at 31%, and subsequent implementation rate at 72%. In^59^, Oncokompas is shown to improve HRQoL and tumor-specific symptom burden, but without any significant effect on secondary outcomes like mental adjustment to cancer.

This study aimed to explore which subgroups of cancer survivors may especially benefit from Oncokompas in terms of HRQoL: it was higher for cancer survivors with low to moderate self-efficacy, high health literacy scores and higher baseline symptom scores^66^. Different outcomes have been reported in^56^, where no significant statistical difference in HRQoL emerged between patients using the app and the control group. Finally, with respect to potential cost savings, no positive effects have been observed by Schuit *et al.*^55^.

In^61^, blended care of group medical consultations and online support for breast cancer follow-up did not improve distress or empowerment, with no statistical differences between intervention and control groups. Decision aids (DA) digital tools showed mixed results: Cuypers *et al.*^63^ reported no significant difference in intervention and control groups of elders patients affected by various types of cancer, whereas Roberto *et al.*^62^ showed how DA for women undergoing cancer screening increased informed choice (without reducing screening participation rate) and awareness about overdiagnosis, and reduced decisional conflict compared to standard brochure (SB). Gorini *et al.*^64^ demonstrated how an interactive empowerment tool (IEm) for breast cancer patients can be used to provide personalized patient profiles and recommendations for physicians, thereby enhancing patient-physician communication, and fostering patients’ empowerment in terms of their participation in the therapeutic process. Giesler *et al.*^65^ reported instead less encouraging results from a web-based randomized control trial on a website presenting patients’ experiences of living with colorectal cancer. Indeed, no effect has been reported at 2 and 6 weeks after baseline on self-efficacy for coping with the disease and on patient competencies, such as coping with emotional distress or dealing with the life-threatening nature of cancer.

Among the digital interventions included in this systematic review, the rate of effectiveness, intended as the capability of such solutions to reach the intended outcomes for which they have been proposed to, varies depending on the area the interventions focuses on: the digital interventions for improving psychophysical well-being proved to be the most effective, with 80% (12 out of 15) of the proposed solutions^19–22,24–26,28,29,32,33^ having a positive impact on mental health; followed by remote monitoring interventions, with 73% (8 out of 11) of included studies^42,44,45,48–53^ resulting useful for patients for real-time self-reporting symptoms and vitals and assessing their quality of life; also digital interventions for managing physical side-effects of cancer treatments are quite effective with 71% (5 out of 7) of the solutions^26,34,35,37,40^ helping coping with physical distress; finally, only the 17% (2 out of 12) solutions^62,64^ proved to enhance empowerment and self-efficacy of cancer patients. Moreover, for the first two categories, the adherence rate is quite high, ranging from 59% to 100%, suggesting a correlation between the effectiveness and the adherence rate of such interventions whose causality could be mutual. However, care should be put in the analysis of these outcomes, whereby small size effect is non negligible in some studies.

## 4. Discussion

### 4.1. Limitations

The results of the studies included in this systematic review have certainly some limitations that need to be taken in consideration in the interpretation and application of the reported findings. A recurring problematic aspect observed across the included studies is a small sample size that limits the statistical power of findings and the applicability of intervention outcomes to broader cancer types or diverse patient populations, hindering the translation of research findings into clinical practice ^20,27,29,44,45,47,57^. This limitation stems from the vast number of cancer types, age range and gender, and is exacerbated by the disproportionate representation of certain demographics within the studies, particularly women with breast cancer and individuals classified as low risk, as these categories have the highest survival rates, and their quality of life allows the participation to experimental studies.

Beyond an insufficient number of participants, another possible source of inaccuracy in the findings stands in the selection criteria, which might yield the risk of biases. Indeed, several works focus on specific subgroups such as specific cancer type patients or stage of treatment (e.g. screening, post-surgical, palliative). On one hand, such a choice may facilitate a deeper understanding of interventions tailored to their needs, but limits the generality of the findings at the same time engendering a risk of demographic bias. In patient inclusion criteria, the requirement of smartphone usage or digital proficiency may exclude individuals who do not possess the necessary digital health literacy, thereby introducing selection bias into the study sample. In particular, older individuals, who may have limited technological literacy or access to digital resources, are potentially underrepresented in studies reliant on internet-based interventions.

Another type of bias may be encountered especially in studies reliant on self-reported measures of health behaviors or psychosocial outcomes^25,43,57,58^, where the participants may tend to provide responses that align with societal norms or expectations, rather than reflecting their true behaviors or experiences. Hence, a suggestion for future investigation is to provide anonymity when possible or, when not possible, to ensure that the interviewers are not the direct practitioners that are treating/have treated the patient to avoid any discomfort.

Finally, most of the studies focus on feasibility, acceptability and short-term effects on the outcome measured to estimate the quality of life of cancer patients, and only few of them investigates the long-term effect with follow-up trials. Moreover, the occurrence of dropouts during follow-up periods^20,24,36,39,46,61^ further exacerbates concerns regarding the representativeness and completeness of the collected data.

Future research should focus more on longitudinal studies able to explore the long-term effects of such initiatives and possibly maximize them. Moreover, the reasons why the participation rate is low should be investigated so to foster participation and to guarantee statistical significance of the results. Finally, the rationale behind dropouts’ rate should be determined towards improving rigor and soundness of the results.

As for the limitations of our review process, we acknowledge that the decision to restrict eligibility to studies in English only, and the search of only three databases may be a source of potential bias in the outcome of our review. In particular, we may have missed relevant initiatives in national languages different from English, whereby not all digital solutions have been published in international journals. In the same vein, focusing only on journal papers whose full-text was available for download, albeit favoring the overall quality of the screened literature, may also have limited the number of potential studies.

### 4.2. Outlook

Albeit the average adoption rate of all the studies included among the target populations is quite low, possibly caused by a low digital literacy (especially among the elders), encouraging levels of adherence are reported, indicating a noteworthy level of usability among participants. Moreover, the consistently high levels of feasibility and satisfaction reported by participants lead to promising outcomes of these interventions, underscoring their perceived viability and acceptability.

With respect to effectiveness, the systematic review reveals mixed results across the examined studies, indicating variability in the achieved outcomes of digital interventions. While some interventions demonstrate promising efficacy in achieving their intended objectives, especially when tackling psychosocial symptoms, others yield non-significant effects. This variability may be attributed to differences in intervention design, target populations, or methodological approaches employed across studies. Nonetheless, despite the mixed findings on effectiveness, the usability of digital interventions consistently emerges as a strength, with participants generally reporting high levels of ease and convenience in utilizing digital resources for cancer support.

A cost reduction for the healthcare system is also a potential strength of digital solutions, with some interventions demonstrating lower costs compared to traditional healthcare while achieving similar effects. However, further exploration is needed to elucidate the cost-effectiveness of digital interventions comprehensively, thereby informing decision-making processes regarding resource allocation and sustainability within EU healthcare systems. Moreover, the systematic review highlights a notable gap in the investigation of long-term effects compared to immediate and short-term follow-ups. While short-term outcomes provide valuable insights into the immediate impact of interventions, understanding the benefits and potential risks associated with continued participation in digital interventions over time is essential for ultimately quantifying their benefits.

Another finding of our systematic literature review is that web-based interventions predominate in the reviewed literature, whereas limited attention is given to screening methodologies and to the integration of wearable devices. Future research should also explore the potential benefits of incorporating diverse intervention modalities, especially considering the ongoing boom of artificial intelligence tools, with the potential of enhancing engagement and effectiveness among cancer patients and their caregivers in EU countries.

Another interesting point that emerges from the literature is that there are mixed attitudes among the healthcare practitioners emerges: for example, van Deursen *et al.*^67^ gathered perspectives of healthcare providers on e-health tools to improve the colorectal cancer care pathway: they highlighted potential opportunities to optimize colorectal cancer care, which, however, may be hindered by limited digital health literacy. Part of the opportunities are related to the partial replacement of in-person care with online services providing patients with information about treatment options or common side effects. The importance of combining personal contact with patients with digital solutions has been underlined, instead, by Slev *et al.*^68^. Indeed, an online focus groups among nurses showed how they value self-management support and e-health for advanced cancer patients but prefer a combination of e-health and personal contact with patients rather a complete substitution of traditional healthcare practices; however, they seem to disregard crucial aspects of self-management, such as self-management support for informal caregivers.

Moreover, a sour note is that, even though the support of informal caregivers is often key for patients undergoing cancer treatment, our search in the literature found only few studies that specifically address the needs and experiences of caregivers within the context of digital interventions, an area that then deserves future research and intervention development. Digital interventions have predominantly been designed with a focus on cancer patients, thereby representing an indirect benefit for caregivers as they enhance patients’ autonomy and alleviate some of the caregiving burden. However, the majority of the interventions are not specifically tailored to address the challenges faced by informal caregivers, such as caregiver strain, mental health issues, and lack of support resources.

Consequently, caregivers may still struggle with significant burdens despite improvements in patients’ autonomy. Future digital interventions must be tailored to address the specific demographic characteristics and cultural contexts of informal caregivers^5^. By doing so, these interventions can better support caregivers’ well-being, ensuring they are adequately equipped to manage their caregiving responsibilities while maintaining their own health and quality of life.

Finally, a thorough evaluation of the digital health solutions should also account for indirect, nonhealthcare related aspects to assess sustainability. Indeed, the fact that most of such initiatives rely on project-specific funding or isolated stakeholders, with limited resources for developmental phases, maintenance, and subsequent enhancements or expansions, poses challenges in maintaining active over a long period the digital interventions, even though they proved to be effective, thereby challenging their sustainability. Thus, a cyclical pattern emerges wherein novel digital solutions are abandoned or face obsolescence due to a lack of sustained updates. At the same time, new initiatives, often duplicating existing designs and functionalities, are introduced independently, perpetuating a cycle of inefficiency and resource redundancy. Ensuring the sustainability of digital interventions for cancer patients necessitates innovative approaches to funding and reimbursement models.

These observations are in line with the Good Practices Guide prepared within the E-health4Cancer project^16^, born from a collaboration between the Greek Cancer Guidance Center Kapa3, the Danish Committee for Health Education, the University of Naples Federico II, and the Greek Carers Network EPIONI, which suggest to design project funding with a specific tapered funding stream allocated for continued use and implementation. One potential suggestion for sustainable funding is to draw inspiration from financial models like the Tobin Tax^69^, wherein a small but specific funding stream is generated, possibly through a general pool. This dedicated funding stream would facilitate the evolution of digital interventions, ensuring a pathway for continuous improvement rather than a series of isolated attempts at innovation. By aligning funding structures with the long-term goals of improving patient outcomes and advancing cancer care, stakeholders can pave the way for meaningful progress in this critical area of healthcare.

## 5. Conclusions

The scope of this systematic literature review was to assess the current state-of-the-art of the academic publications on digital solutions for the support of cancer patients and their caregivers across the EU countries. Exploring the interventions proposed in the last decade reveals a substantial interest towards these news tools, as evidenced by the large number of scientific articles featuring various study design. We found scientific publications to be heterogeneous across EU countries and cancer types, with a prevalence of articles from the Netherlands and with a user base of breast cancer patients, in line with the highest incidence and survival rates in Europe, respectively. Then, we clustered the proposed digital interventions according to the main themes onto which they focus on: (a) psychophysical well-being; (b) reduction of physical cancer treatments side effect; (c) remote monitoring; (d) empowerment and self-efficacy. Finally, we found that the most effective solutions are those proposed to enhance mental health and psychological issues, followed by those focusing on remote monitoring. Overall, the review underlines the great interest and potential of these digital tools that hopefully will be integrated in the daily basis routine for cancer patients and will be extended also to their caregivers. Indeed, the outcomes reported in the selected studies generally show a higher effectiveness of cancer care thanks to digital solutions. Moreover, even when the overall patient quality of life was not significantly improved compared to traditional solutions, additional benefits need to be considered in terms of privacy, cost/benefit ratio, and adherence to treatment.

## CRediT author statement

**Camilla Ancona:** Conceptualization, Methodology, Investigation, Data Curation, Writing – Original Draft preparation, Visualization. **Emanuele Caroppo:** Conceptualization, Writing-Original draft preparation, Supervision. **Pietro De Lellis:** Conceptualization, Methodology, Investigation, Writing – Original Draft preparation, Supervision.

## Data availability

Data are available upon reasonable request to the corresponding author.

## Acknowledgements

These authors wish to acknowledge the E-Health4Cancer (project code: 2022-2-EL01-KA210-ADU-000097120) project funded by the European Union under the Erasmus+ KA210-ADU program, in cooperation between the University of Naples Federico II, the Danish Committee for Health Education (DCHE), the Greek Carers Network - EPIONI and the Cancer Guidance Center - Kapa3 (coordinator of the project).

